# Prostate cancer risk stratification improved across multiple ancestries with new polygenic hazard score

**DOI:** 10.1101/2021.08.14.21261931

**Authors:** Minh-Phuong Huynh-Le, Roshan Karunamuni, Chun Chieh Fan, Lui Asona, Wesley K Thompson, Maria Elena Martinez, Rosalind A Eeles, Zsofia Kote-Jarai, Kenneth R Muir, Artitaya Lophatananon, Johanna Schleutker, Nora Pashayan, Jyotsna Batra, Henrik Grönberg, David E Neal, Børge G Nordestgaard, Catherine M Tangen, Robert J MacInnis, Alicja Wolk, Demetrius Albanes, Christopher A Haiman, Ruth C Travis, William J Blot, Janet L Stanford, Lorelei A Mucci, Catharine M L West, Sune F Nielsen, Adam S Kibel, Olivier Cussenot, Sonja I Berndt, Stella Koutros, Karina Dalsgaard Sørensen, Cezary Cybulski, Eli Marie Grindedal, Florence Menegaux, Jong Y Park, Sue A Ingles, Christiane Maier, Robert J Hamilton, Barry S Rosenstein, Yong-Jie Lu, Stephen Watya, Ana Vega, Manolis Kogevinas, Fredrik Wiklund, Kathryn L Penney, Chad D Huff, Manuel R Teixeira, Luc Multigner, Robin J Leach, Hermann Brenner, Esther M John, Radka Kaneva, Christopher J Logothetis, Susan L Neuhausen, Kim De Ruyck, Piet Ost, Azad Razack, Lisa F Newcomb, Jay H Fowke, Marija Gamulin, Aswin Abraham, Frank Claessens, Jose Esteban Castelao, Paul A Townsend, Dana C Crawford, Gyorgy Petrovics, Ron HN van Schaik, Marie-Élise Parent, Jennifer J Hu, Wei Zheng, Ian G Mills, Ole A Andreassen, Anders M Dale, Tyler M Seibert, UKGPCS collaborators, APCB (Australian Prostate Cancer BioResource), NC-LA PCaP Investigators, The IMPACT Study Steering Committee and Collaborators, Canary PASS Investigators, The Profile Study Steering Committee, The PRACTICAL Consortium

**Author notes:** **Corresponding Author:** Tyler M. Seibert, MD, PhD, Assistant Professor, Department of Radiation Medicine and Applied Sciences, Department of Radiology, Department of Bioengineering, Center for Multimodal Imaging and Genetics, University of California San Diego, 9500 Gilman Dr. Mail Code 0861, La Jolla, CA 92093-0861. http://www.icr.ac.uk/our-research/research-divisions/division-of-genetics-and-epidemiology/oncogenetics/research-projects/ukgpcs/ukgpcs-collaborators. https://pcap.bioinf.unc.edu/ (NC-LA PCaP Investigators). http://impact.icr.ac.uk (The IMPACT Study Steering Committee and Collaborators). http://www.cancerresearchuk.org/about-cancer/find-a-clinical-trial/a-study-find-out-looking-gene-changes-would-be-useful-in-screening-for-prostate-cancer-profile-pilot (The Profile Study Steering Committee). Prostate Cancer Association Group to Investigate Cancer Associated Alterations in the Genome Consortium (PRACTICAL, http://practical.icr.ac.uk/). Additional members of the consortium are provided in the Supplementary Information. **Competing interests:** All authors declare no support from any organization for the submitted work except as follows: AMD and TMS report a past research grant from the US Department of Defense. OAA reports research grants from K.G Jebsen Stiftelsen, Research Council of Norway, and South East Norway Health Authority. Authors declare no financial relationships with any organizations that might have an interest in the submitted work in the previous three years except as follows, with all of these relationships outside the present study: TMS reports honoraria from Multimodal Imaging Services Corporation, Varian Medical Systems, and WebMD; he has an equity interest in CorTechs Labs and also serves on its Scientific Advisory Board. These companies might potentially benefit from the research results. The terms of this arrangement have been reviewed and approved by the University of California San Diego in accordance with its conflict-of-interest policies. OAA reports speaker honoraria from Lundbeck. Authors declare no other relationships or activities that could appear to have influenced the submitted work except as follows: OAA has a patent application # U. S. 20150356243 pending; AMD also applied for this patent application and assigned it to UC San Diego. AMD has additional disclosures outside the present work: founder, equity holder, and advisory board member for CorTechs Labs, Inc.; founder and equity holder in HealthLytix, Inc., advisory board member of Human Longevity, Inc.; recipient of nonfinancial research support from General Electric Healthcare. OAA is a consultant for HealthLytix, Inc. Additional acknowledgments for the PRACTICAL consortium and contributing studies are described in the Supplementary Information. **Ethics statement:** The present analyses used de-identified data from the PRACTICAL consortium and have been approved by the Institutional Review Board at the University of California San Diego. All contributing studies were approved by the relevant ethics committees and performed in accordance with the Declaration of Helsinki.

## Abstract

**Introduction:** Prostate cancer risk stratification using single-nucleotide polymorphisms (SNPs) demonstrates considerable promise in men of European, Asian, and African genetic ancestries, but there is still need for increased accuracy. We evaluated whether including additional SNPs in a prostate cancer polygenic hazard score (PHS) would improve associations with clinically significant prostate cancer in multi-ancestry datasets.

**Methods:** In total, 299 SNPs previously associated with prostate cancer were evaluated for inclusion in a new PHS, using a LASSO-regularized Cox proportional hazards model in a training dataset of 72,181 men from the PRACTICAL Consortium. The PHS model was evaluated in four testing datasets: African ancestry, Asian ancestry, and two of European Ancestry—the Cohort of Swedish Men (COSM) and the ProtecT study. Hazard ratios (HRs) were estimated to compare men with high versus low PHS for association with clinically significant, with any, and with fatal prostate cancer. The impact of genetic risk stratification on the positive predictive value (PPV) of PSA testing for clinically significant prostate cancer was also measured.

**Results:** The final model (PHS290) had 290 SNPs with non-zero coefficients. Comparing, for example, the highest and lowest quintiles of PHS290, the hazard ratios (HRs) for clinically significant prostate cancer were 13.73 [95% CI: 12.43-15.16] in ProtecT, 7.07 [6.58-7.60] in African ancestry, 10.31 [9.58-11.11] in Asian ancestry, and 11.18 [10.34-12.09] in COSM. Similar results were seen for association with any and fatal prostate cancer. Without PHS stratification, the PPV of PSA testing for clinically significant prostate cancer in ProtecT was 0.12 (0.11-0.14). For the top 20% and top 5% of PHS290, the PPV was 0.19 (0.15-0.22) and 0.26 (0.19-0.33), respectively.

**Conclusion:** We demonstrate better genetic risk stratification for clinically significant prostate cancer than prior versions of PHS in multi-ancestry datasets. This is promising for implementing precision-medicine approaches to prostate cancer screening decisions in diverse populations.

## Introduction

Identification of men at greatest risk of developing prostate cancer remains an important challenge. Risk stratification using common genetic markers, such as single-nucleotide polymorphisms (SNPs), shows promise toward more effectively identifying men at greatest risk of developing aggressive or fatal prostate cancer^1,2^. Analyses of benefit, harm, and cost-effectiveness support use of genomic risk stratification to guide prostate cancer screening^3,4^. Ensuring these tools perform well in diverse populations is important to ensure risk stratification is optimal for all men and avoid exacerbating existing health disparities^5,6^.

We previously developed a polygenic hazard score (PHS) and demonstrated in an independent European dataset that the PHS was associated with age at diagnosis of clinically significant prostate cancer^1^. Risk stratification with this score also improved the accuracy of PSA testing^7,8^. We then validated the PHS model (with 46 SNPs) for association with age at diagnosis and with prostate-cancer specific mortality in a dataset that included men of diverse descent, including European, African, and Asian ancestry^2^. We have also improved performance in men of African ancestry by searching for SNPs within that subpopulation^9,10^.

Recent meta-analyses have identified over 200 SNPs associated with prostate cancer, including some identified through subset analyses in men of non-European ancestry^11,12^. Given the increasing number of SNPs associated with prostate cancer, we evaluated whether including more SNPs in a prostate cancer PHS would improve associations with clinically significant prostate cancer in multi-ancestry datasets.

## Methods

### Participants

Genotype and phenotype data, all de-identified, were obtained from the PRACTICAL consortium. Participants had previously been genotyped via the OncoArray^13^ or the iCOGs^14^ chips; 90,638 men were available for this analysis.

The available data were split into a training dataset and four testing datasets, taking into account prior power analyses and PHS association results^7,15,16^. The training dataset for the model included 72,181 men of European genetic ancestry genotyped via OncoArray (24,010 controls and 48,171 cases). The four testing datasets included: 1) men of African ancestry (n=6,253: 3,013 controls and 3,240 cases), 2) men of Asian ancestry (n=2,378: 1,184 controls and 1,194 cases), 3) the Cohort of Swedish Men (COSM) population-based cohort with long-term outcomes^17^ (n=3,279: 1,116 controls, 2,163 cases, and 278 prostate cancer deaths), and 4) the ProtecT population-based prospective trial with screening (prostate-specific antigen, PSA) and biopsy outcomes for both cases and controls (n=6,411: 4,828 controls and 1,583 cases)^18^.

### Polygenic Hazard Score Model Development using LASSO regularization

We sought to develop an optimal, integrated PHS model with candidate SNPs chosen from those used in the prior PHS and from those identified as susceptibility loci for prostate cancer in a genome-wide trans-ancestry meta-analysis^11^. Candidate SNPs included the 46 from the original PHS development and 269 from the meta-analysis. A machine-learning, LASSO-regularized Cox proportional hazards model approach was used to objectively select SNPs and estimate weights, as described previously^8^.

There were 299 unique candidate SNPs associated with prostate cancer consistent across the training and testing datasets used in the present work. We first identified SNP pairs (among the 299 candidates) that were highly correlated (*r*^2^>0.95). Each of these paired, correlated SNPs was tested in a univariable Cox proportional hazards model for association with age at prostate cancer diagnosis; the SNP with the larger *p*-value was eliminated from inclusion in the model. All other (unpaired) SNPs were included as candidates for the present PHS model.

The *R* (version 4.0.1) “glmnet” package was used to estimate the LASSO-regularized Cox proportional hazards model^19,20^. Age at prostate cancer diagnosis was the time to event, and the predictor variables included the genotype allele counts of candidate SNPs and first four European ancestry principal components. Controls were censored at age of last follow-up. The LASSO-regularized model’s hyper-parameter (lambda) was selected using 10-fold cross-validation^19,20^. The final form of the LASSO model was estimated using the lambda value that minimized the mean cross-validated error.

### Association with Prostate Cancer

We evaluated the association of the adapted PHS with clinically significant prostate cancer, as well as any prostate cancer, via Cox proportional hazards models in each of the four testing datasets. Clinically significant prostate cancer was defined to be a prostate cancer case with Gleason score ≥7, PSA ≥10 ng/mL, T3-T4 stage, nodal metastases, or distant metastases^21^.

As the COSM dataset had long-term follow up data available^17^, we additionally evaluated the adapted PHS for association with age at prostate cancer death^16^. There were 278 deaths from prostate cancer in the COSM dataset.

### Hazard Ratio Performance

Hazard ratios between the top 5% and middle 40% (HR_95/50_), top 20% and middle 40% (_HR80/50_), bottom 20% and middle 40% (HR_20/50_), and top and bottom 20% (HR_80/20_) were estimated for any, for clinically significant, and for fatal prostate cancer. Percentiles of genetic risk were determined within the controls the training set with age less than 70 years^1,2,7,8^.

### Family History

Given that family history of prostate cancer is currently one of the most useful clinical risk factors for the development of prostate cancer, we used Cox proportional hazards models to assess family history for association with any, with clinically significant, or with fatal prostate cancer. Family history of prostate cancer was defined as presence or absence of a first-degree relative diagnosed with prostate cancer. Multivariable models using both family history and the adapted PHS were compared to using family history alone via a log-likelihood test with α=0.01. HRs were calculated for each variable: HRs for PHS in the multivariable models were estimated as the HR_80/20_ in each testing dataset (e.g., men in the highest vs. lowest quintile of genetic risk by PHS). HRs for family history of prostate cancer were estimated as the exponent of the beta from the multivariable Cox regression. As done previously^1,7^, *p*-values were truncated at <10^−16^.

### Positive Predictive Value Performance

Positive predictive value (PPV) performance of PSA testing was calculated using data from the population-based ProtecT screening study^18^ (prostate biopsy results were available for both cases and controls with a positive PSA [≥3 ng/mL]). To estimate the PPV and confidence intervals, we generated 1,000 bootstrap samples using ProtecT participants with positive PSA, while maintaining the 1:2 case:control ratio in the ProtecT dataset. PPV was calculated as the proportion of PSA-positive participants who were diagnosed with clinically significant prostate cancer on biopsy, looking at those participants in the top 5 (PPV_95_) or top 20 percentiles (PPV_80_) of PHS genetic risk.

### Cumulative incidence curves for PHS

Genetic-risk-stratified cumulative incidence curves for prostate cancer were derived using previously described methods^7,8^. Briefly, age-specific population data from the United Kingdom (Cancer Research UK^7^) were used to estimate prostate cancer incidence for men aged 40-70 years. Data from the population-based Cluster Randomized Trial of PSA Testing for Prostate Cancer (CAP) trial^7^ were used to adjust this population incidence curve to reflect the age-specific cumulative incidence of clinically significant and non-clinically-significant prostate cancer. Genetic-risk-stratified cumulative incidence curves were then calculated for men in the upper 5 and 20 percentiles of PHS genetic risk by multiplying the prostate-cancer-specific cumulative incidence by the mean value of HR_95/50_ and HR_80/50_ in the testing dataset, respectively.

## Results

A total of 290 SNPs returned non-zero SNP coefficients using the regularization-weight selection and were included in the final model, called PHS290 (**Supplementary Material**).

HR performance of PHS290 demonstrates risk stratification across percentiles of genetic risk (**Table 1**). Comparing the top and bottom quintiles of genetic risk for clinically significant prostate cancer, men with high PHS had HRs of 13.73 [12.43-15.16], 7.07 [6.58-7.60], 10.31 [9.58-11.11], and 11.18 [10.34-12.09] in the ProtecT, African, Asian, and COSM datasets, respectively. Similar risk stratification was seen when evaluating risk of any prostate cancer. Finally, when comparing the top and bottom quintiles of genetic risk in the COSM dataset, men with high PHS had a HR of 7.73 [6.45-9.27] for prostate cancer death.

**Table 1.** Hazard ratio (HR) performance in the four testing datasets. HRs are shown with mean sample-weight-corrected values and 95% confidence intervals. Calculations were done using age at diagnosis of any or of clinically significant prostate cancer across all datasets, respectively, and also with age at prostate cancer death for the COSM dataset.

| Dataset | Prostate Cancer Type | HRs and 95% CI |  |  |  |
| --- | --- | --- | --- | --- | --- |
|  |  | HR<sub>20/50</sub> | HR<sub>80/50</sub> | HR<sub>95/50</sub> | HR<sub>80/20</sub> |
| ProtecT<br>n=6,411 | Any | 0.31 [0.30-0.32] | 3.47 [3.36-3.58] | 5.93 [5.67-6.21] | 11.16 [10.48-11.88] |
|  | Clinically significant | 0.28 [0.27-0.30] | 3.86 [3.67-4.06] | 6.91 [6.42-7.44] | 13.73 [12.43-15.16] |
| African<br>n=6,253 | Any | 0.43 [0.42-0.45] | 2.58 [2.50-2.67] | 3.69 [3.52-3.86] | 5.95 [5.59-6.34] |
|  | Clinically significant | 0.40 [0.39-0.41] | 2.82 [2.72-2.93] | 4.33 [4.10-4.57] | 7.07 [6.58-7.60] |
| Asian<br>n=2,378 | Any | 0.34 [0.33-0.35] | 2.99 [2.89-3.08] | 5.08 [4.85-5.33] | 8.75 [8.21-9.32] |
|  | Clinically significant | 0.31 [0.30-0.33] | 3.24 [3.12-3.36] | 5.66 [5.46-5.98] | 10.31 [9.58-11.11] |
| COSM<br>n=3,279 | Any | 0.33 [0.32-0.34] | 3.54 [3.42-3.65] | 5.77 [5.51-6.04] | 10.87 [10.21-11.57] |
|  | Clinically significant | 0.32 [0.31-0.33] | 3.59 [3.45-3.75] | 5.91 [5.58-6.26] | 11.18 [10.34-12.09] |
|  | Fatal | 0.38 [0.35-0.42] | 2.95 [2.68-3.25] | 4.49 [3.93-5.13] | 7.73 [6.45-9.27] |

### Family history and PHS290

The combination of family history and PHS performed better than family history, alone, for clinically significant prostate cancer (and for any prostate cancer) in each of the four testing datasets (log-likelihood *p*<10^−16^; **Table 2**). Additionally, family history and PHS together performed better than family history, alone, for fatal prostate cancer in the COSM dataset (log-likelihood *p*<^10-16^).

**Table 2.** Multivariable Cox models with both PHS and family history of prostate cancer (defined as ≥1 first-degree relative affected) for association with any prostate cancer, with clinically significant prostate cancer, and with fatal prostate cancer. Analyses were limited to participants with known family history. Beta and *z*-scores refer to the overall association (within the multivariable Cox regression) with the endpoint of interest, within the corresponding testing dataset. The *p*-values reported are two-tailed from the multivariable Cox models.

| Dataset | Variable | Any Prostate Cancer |  |  |  |
| --- | --- | --- | --- | --- | --- |
|  |  | beta | z-score | p-value | HR |
| ProtecT (iCOGs),<br>n=5,703 | PHS | 2.11 | 72.1 | $<10^{-16}$ | 8.13 |
|  | Family history | 0.06 | 1.7 | 0.1 | 1.07 |
| African (OncoArray),<br>n=5,557 | PHS | 1.64 | 53.8 | $<10^{-16}$ | 5.03 |
| | Family history | 1.13 | 46.6 | $<10^{-16}$ | 3.09 |
| Asian (OncoArray);<br>n=1,028 | PHS | 1.99 | 68.5 | $<10^{-16}$ | 7.64 |
| | Family history | 0.45 | 13.1 | $<10^{-16}$ | 1.56 |
| COSM (OncoArray),<br>n=2,453 | PHS | 2.13 | 71.2 | $<10^{-16}$ | 8.71 |
| | Family history | 0.53 | 19.0 | $<10^{-16}$ | 1.70 |
| Dataset | Variable | Clinically Significant Prostate Cancer |  |  |  |
|  |  | beta | z-score | p-value | HR |
| ProtecT (iCOGs),<br>n=5,703 | PHS | 2.32 | 49.8 | $<10^{-16}$ | 10.01 |
|  | Family history | -0.01 | -0.2 | 0.82 | 0.99 |
| African (OncoArray),<br>n=5,557 | PHS | 1.67 | 37.4 | $<10^{-16}$ | 5.19 |
| | Family history | 1.17 | 33.2 | $<10^{-16}$ | 3.22 |
| Asian (OncoArray),<br>n=1,028 | PHS | 1.89 | 50.4 | $<10^{-16}$ | 6.90 |
|  | Family history | 0.13 | 2.5 | 0.012 | 1.14 |
| COSM (OncoArray),<br>n=2,453 | PHS | 2.13 | 56.9 | $<10^{-16}$ | 6.90 |
| | Family history | 0.45 | 12.6 | $<10^{-16}$ | 1.57 |
| Dataset | Variable | Fatal Prostate Cancer |  |  |  |
|  |  | beta | z-score | p-value | HR |
| COSM (OncoArray),<br>n=2,453 | PHS | 1.68 | 19.8 | $<10^{-16}$ | 5.51 |
| | Family history | 0.43 | 5.0 | $4.7 \times 10^{-7}$ | 1.53 |

### Positive predictive value performance of PHS290

The PPV of PSA testing for clinically significant prostate cancer was 0.19 (0.15-0.22) for the top 20% of genetic risk (PPV_80_) and 0.26 (0.19-0.33) for the top 5% of genetic risk (PPV_80_; **Figure 1**). Both were greater than the overall PPV of PSA alone, which was 0.12 (0.11-0.14).

**Figure 1.**
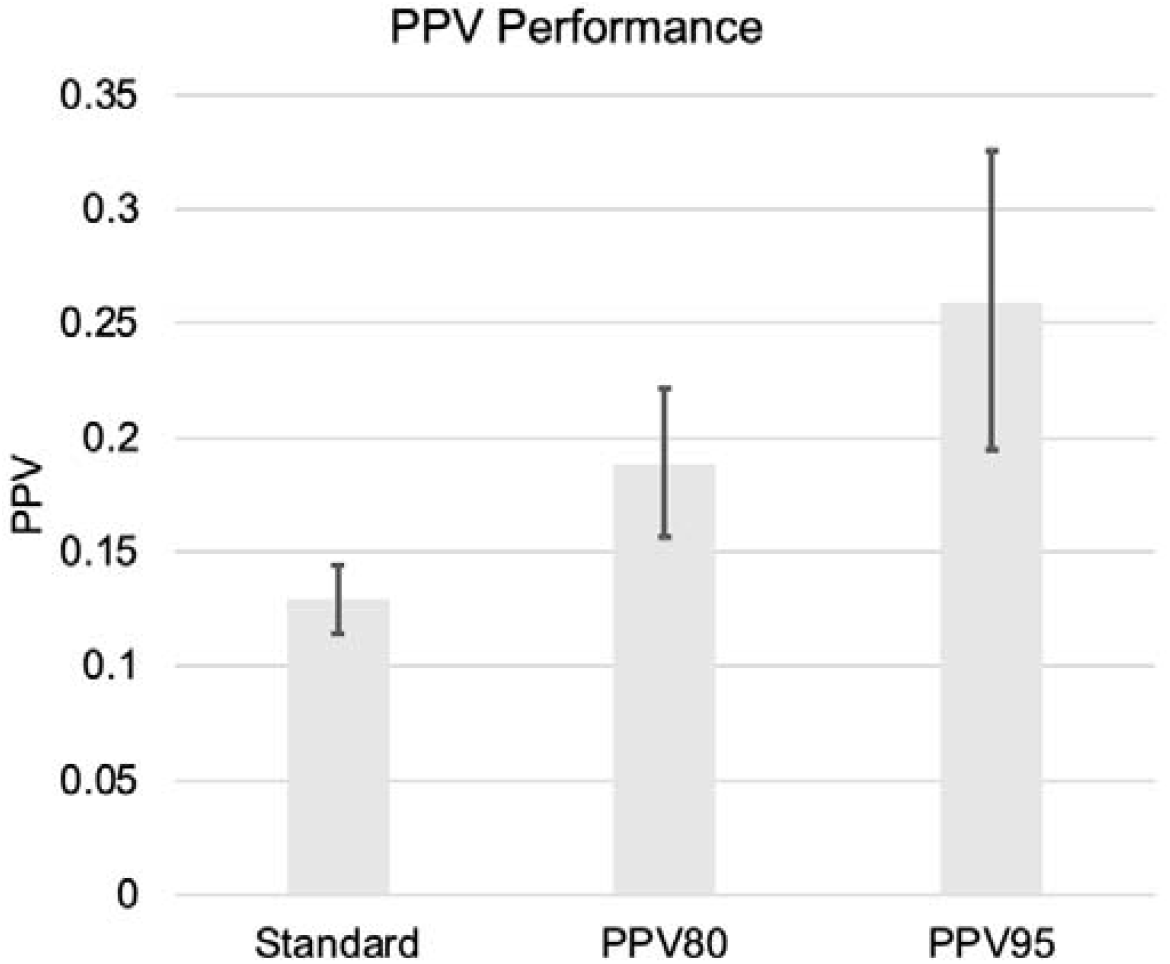
PPV performance in the ProtecT dataset for clinically significant prostate cancer, estimated using 3 approaches: standard (not using PHS), top 20% of PHS values (PPV80), and top 5% of PHS values (PPV95). Error bars are 95% bootstrap confidence intervals.

### Cumulative incidence curves for PHS290

Genetic-risk-stratified cumulative incidence curves for clinically significant and non-clinically significant prostate cancer demonstrate greater prostate cancer incidence with higher genetic-adjusted risk (**Figure 2**).

**Figure 2.**
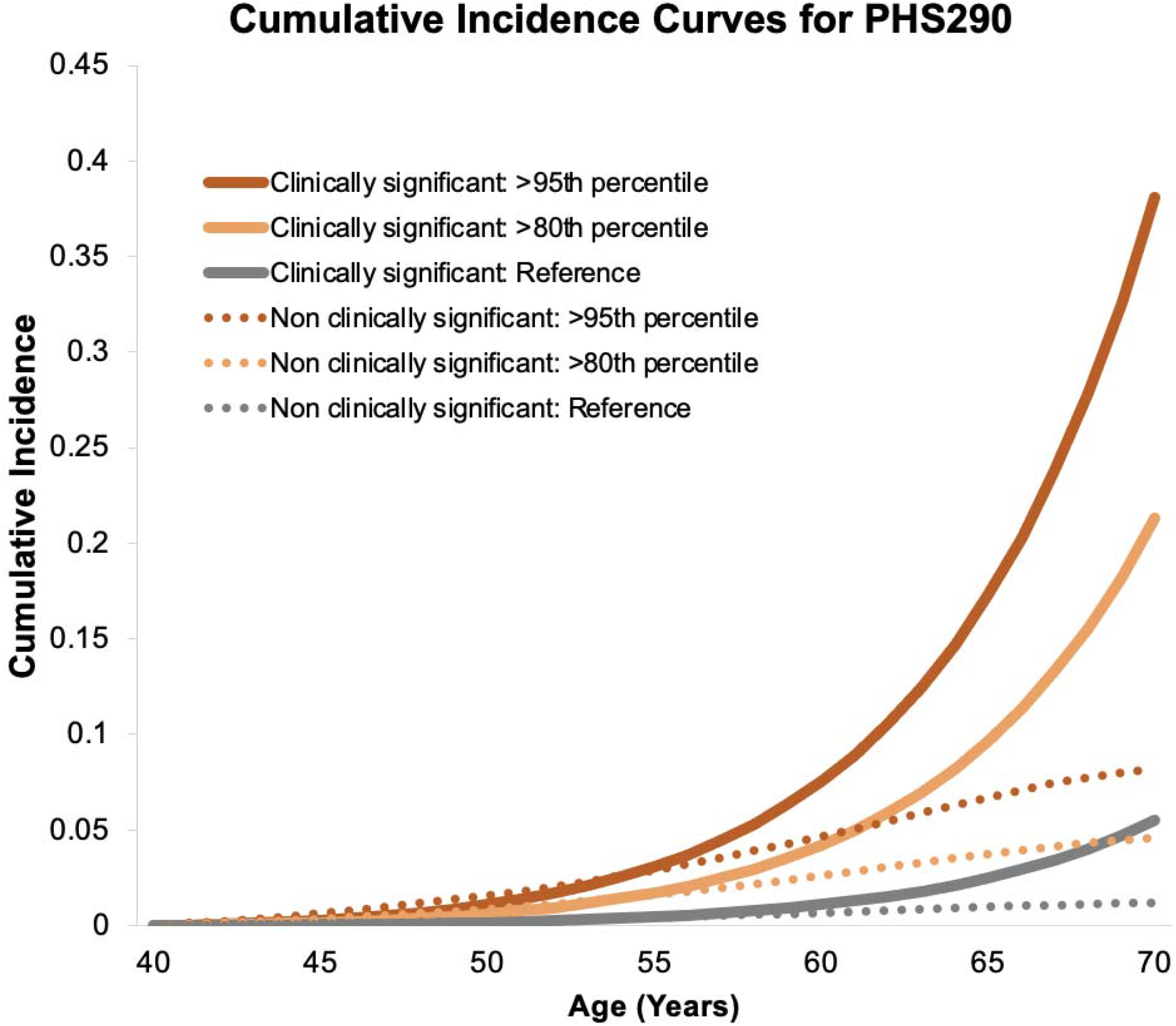
Genetic-risk-adjusted cumulative incidence curves for PHS290. Curves are shown for the upper 5^th^ (>95^th^) and upper 20^th^ (>80^th^) percentile of PHS290 for clinically significant and non-clinically-significant prostate cancer. The reference curves represent the overall population average from the UK.

## Discussion

The improved PHS (PHS290) demonstrates excellent genetic risk stratification, including for clinically significant prostate cancer. This was true in four separate testing sets of varied genetic ancestry (Asian, African, European). Additionally, PHS290 was associated with lifetime prostate-cancer-specific mortality in a population-based cohort^16^. Hazard ratios with PHS290 are larger for each of these associations than those reported for previous versions of PHS^2,8^, demonstrating the value of incorporating SNPs from genome-wide meta-analysis and fine-mapping. The improvements demonstrated here are promising for implementing personalized approaches to prostate cancer screening decisions in diverse populations.

Health disparities are a major problem in prostate cancer. Given the exclusion of non-European data in most genome studies^5,22–24^, it is important that the pool of candidate SNPs here included those identified from a recent trans-ancestry meta-analysis^11^. Testing and improving performance of genomic risk scores in diverse populations is critical to equitable implementation of these new tools and important for avoiding exacerbation of existing disparities. PHS290 still performs better in men of European genetic ancestry—an expected result, given the much greater data availability in that population. Further genomic studies in diverse populations are essential, as diversity in model development improves performance in diverse populations^9,10^.

The intersection of social constructs like race/ethnicity and genomics also raises interesting and entangled challenges. Even availability of genomic data is only part of the problem, as disparities in health outcomes are rooted in systemic racism and inequities in access to healthcare^25,26^. Genotypic ancestry may be a step toward biology, but the continental groups still represent an oversimplification of genetic diversity and a pre-determined assumption that socially defined categories have biological meaning in all contexts. We have previously shown that agnostic genetic clusters are informative for subgroup analyses^2^, and this approach may be a better way forward, provided the genomic diversity of the whole population is represented in the available data. Here, we have used genotypic ancestry to evaluate the potential differential performance in groups historically excluded from large-scale genomic studies. We also note that local ancestry may be a critical consideration in admixed populations. We have found previously that PHS performance can vary by the makeup of a *region* of the genome, beyond what is explained by *global* ancestry categories assigned for an individual’s entire genome^10^. Despite these challenges and opportunities for future improvement, the current results demonstrate PHS290 does provide meaningful risk stratification in diverse datasets.

The cumulative incidence of clinically significant prostate cancer is heavily influenced by age-specific genetic risk, as demonstrated by genetic-risk-stratified cumulative incidence curves (**Figure 2**). As men with high PHS290 age, the incidence curve for clinically significant prostate cancer increases dramatically, prominently separating from the incidence for non-clinically-significant prostate cancer. This effect is driven by the high HR for clinically significant prostate cancer in these men, combined with increasing incidence specifically of more clinically significant cancers as men age^7,27^. Furthermore, we found that risk stratification with PHS290 improved accuracy of PSA testing, as assessed by probability of a positive PSA test leading to a diagnosis of clinically significant cancer on biopsy. Consistent with a prior study^8^, this improvement in PPV of PSA testing was not better when using PHS290 than when using PHS46 in this dataset^2^. PPV analyses in larger datasets could permit finer granularity for age-specific genetic risk to assess whether the increased HRs of PHS290 might translate to better performance of PSA testing than that achieved already with PHS46.

The HRs reported here suggest clinical relevance for PHS290. Predictive tools in routine clinical use for other diseases (e.g., breast cancer, diabetes, and cardiovascular disease) have reported HRs of approximately 1-3 for endpoints of interest^28–31^. Current guidelines recommend earlier and more frequent consideration of prostate cancer screening for men with a family history of prostate cancer or African ancestry, citing an elevated risk 28-80% above that of men without these risk factors^32–34^. Guidelines more strongly recommend earlier and more frequent screening in men with germline mutations in *BRCA2*, which are rare but are estimated to infer up to 3-fold increased risk^33–35^. In the present study, men in the top 20% for PHS290 (compared to men with *average* risk) had HRs for clinically significant prostate cancer of approximately 2.8 to 3.9. For men in the top 5% for PHS290, those HRs increase to 4.3 to 6.9, depending on ancestry. While individuals with high polygenic risk may also develop low-grade prostate cancer in their lifetime, the time-to-event analysis applied here shows that high genetic risk confers a greater hazard for prostate cancer death. This finding is consistent with prior reports, though the effect size is larger with PHS290^2,16,33^.

Our work has some limitations. First, the weights were calculated in men of European genetic ancestry alone, although SNP candidate selection was performed in multi-ancestry analyses. Future studies evaluating PHS for prostate cancer risk stratification will include non-Europeans in SNP weight calculations. The available data did not permit testing of PPV or association with fatal disease in non-European populations. Moreover, the cumulative incidence curves here are specific to the UK, where we had the most robust population age-specific incidence data for clinically significant prostate cancer. The testing sets used in the present study did represent a very small proportion of the data used for candidate SNP identification in the prior genome-wide association meta-analysis (as opposed to the development of PHS46, which was performed in a dataset completely independent of the validation dataset) ^2 11^. However, the training and testing sets were kept separate in the present study, and the use of the LASSO-regularized Cox model reduces over-fitting^36^.

The PHS290 described here has the strongest reported association with prostate cancer in men of European, African, and Asian genetic ancestry. The score was also associated with lifetime prostate-cancer-specific mortality in a population-based cohort. A performance gap remains between genetic ancestry groups that might be closed through development using more data from men of Asian or African ancestry. Nonetheless, the results here suggest PHS290 may improve prostate cancer risk-stratification efforts in multi-ancestry populations.

## Data Availability

Prostate Cancer Association Group to Investigate Cancer Associated Alterations in the Genome (PRACTICAL) Consortium data are available upon request to the Data Access Committee (http://practical.icr.ac.uk). Questions and requests for further information may be directed to.

