## Supplementary Information for "Prostate cancer risk stratification improved across multiple ancestries with new polygenic hazard score"

**Supplementary Table 1**. SNP identifier, chromosome, effect allele, reference allele, and position (based on version 37), and beta (model weight) for the 290 SNPs used in the calculation of PHS290. The effect-allele-frequency (EAF) was estimated from controls in the training dataset with age less than 70 years. The HGNC identifier and Variant Consequence for each SNP were extracted from dbSNP.

| **SNP ID** | **Chr** | **Effect** | **Ref** | **Position** | **beta** | **EAF** | **HGNC** | **Variant Consequence** |
| --- | --- | --- | --- | --- | --- | --- | --- | --- |
| rs12262998 | 10 | C | T | 104428716 | 0.0254 | 0.6792 |  |  |
| rs10885396 | 10 | T | C | 114711755 | 0.0095 | 0.5434 | TCF7L2 | intron_variant |
| rs4558107 | 10 | A | G | 122794926 | 0.0227 | 0.3932 |  |  |
| rs140783917 | 10 | C | T | 122834482 | 0.1275 | 0.9989 |  |  |
| rs10788167 | 10 | T | A | 123054018 | 0.0249 | 0.7674 |  |  |
| rs10749415 | 10 | A | G | 123185303 | 0.0602 | 0.9469 |  |  |
| rs12769682 | 10 | C | G | 126697494 | 0.0358 | 0.2720 | CTBP2 | intron_variant;non_coding_transcript_variant |
| rs7075427 | 10 | A | C | 46104943 | 0.0069 | 0.9209 |  |  |
| rs11599847 | 10 | T | C | 47599029 | -0.0171 | 0.9629 |  |  |
| rs10993994 | 10 | T | C | 51549496 | 0.1176 | 0.3906 |  |  |
| rs11817544 | 10 | C | A | 80236999 | 0.0338 | 0.9413 | LINC00856 | non_coding_transcript_variant;intron_variant |
| rs12412705 | 10 | C | T | 80835998 | 0.0180 | 0.0692 | ZMIZ1 | intron_variant |
| rs12781100 | 10 | T | C | 838636 | 0.0543 | 0.1600 |  |  |
| rs1935581 | 10 | C | T | 90195149 | 0.0323 | 0.6297 | RNLS | intron_variant;non_coding_transcript_variant |
| rs11568818 | 11 | T | C | 102401661 | 0.0333 | 0.5494 |  |  |
| rs74911261 | 11 | A | G | 108357137 | 0.0598 | 0.0245 | KDELC2 | missense_variant;NMD_transcript_variant;non_coding_transcript_exon_variant |
| rs138466039 | 11 | T | C | 125054793 | 0.0801 | 0.0103 | PKNOX2 | intron_variant;non_coding_transcript_variant;NMD_transcript_variant |
| rs878987 | 11 | G | A | 134266372 | 0.0241 | 0.1460 | B3GAT1 | intron_variant;non_coding_transcript_variant |
| rs1881502 | 11 | T | C | 1507512 | 0.0107 | 0.1903 | MOB2 | intron_variant;non_coding_transcript_variant |
| rs72853963 | 11 | A | G | 2224664 | 0.0190 | 0.1822 |  |  |
| rs11043143 | 11 | T | C | 2234093 | 0.0735 | 0.1948 |  |  |
| rs68010938 | 11 | T | TA | 47428209 | 0.0186 | 0.3047 |  | non_coding_transcript_variant;intron_variant |
| rs1048374 | 11 | G | A | 58902679 | -0.0747 | 0.0037 |  | non_coding_transcript_exon_variant |
| rs2277283 | 11 | C | T | 61908440 | 0.0356 | 0.3045 | INCENP | missense_variant;non_coding_transcript_exon_variant |
| rs12785905 | 11 | C | G | 66951965 | 0.0247 | 0.0389 | KDM2A | intron_variant |
| rs3018690 | 11 | T | C | 68882926 | 0.0297 | 0.4431 |  |  |
| rs11825796 | 11 | A | G | 68980788 | -0.0164 | 0.2625 |  |  |
| rs12275055 | 11 | G | A | 68981359 | 0.0453 | 0.1652 |  |  |
| chr11_68985583 | 11 | C | T | 68985583 | -0.0531 | 0.4928 |  |  |
| rs11228580 | 11 | C | T | 69002342 | 0.0094 | 0.1655 |  |  |
| rs3918298 | 11 | A | G | 69463273 | 0.0824 | 0.0274 | CCND1 | intron_variant;non_coding_transcript_variant |
| rs61890184 | 11 | A | G | 7547587 | 0.0334 | 0.1179 | PPFIBP2 | intron_variant |
| rs56159348 | 11 | T | G | 76267331 | 0.0259 | 0.6783 |  |  |
| rs77121786 | 12 | G | T | 102446675 | 0.0341 | 0.1981 | CCDC53 | NMD_transcript_variant;intron_variant;non_coding_transcript_variant |
| rs1270884 | 12 | A | G | 114685571 | 0.0239 | 0.4816 |  |  |
| rs2066827 | 12 | T | G | 12871099 | 0.0348 | 0.7603 | CDKN1B | non_coding_transcript_variant;intron_variant;missense_variant;non_coding_transcript_variant;intron_variant;coding_sequence_variant |
| rs77216612 | 12 | A | G | 12877983 | 0.0030 | 0.7240 |  |  |
| rs7295014 | 12 | G | A | 133067989 | 0.0243 | 0.3349 | FBRSL1 | intron_variant |
| rs10845938 | 12 | G | A | 14416918 | 0.0315 | 0.5489 |  |  |
| rs80130819 | 12 | A | C | 48419618 | 0.0519 | 0.9104 |  |  |
| rs56222401 | 12 | G | A | 49672714 | 0.0180 | 0.2499 |  |  |
| rs10875943 | 12 | C | T | 49676010 | 0.0178 | 0.2810 |  |  |
| rs113925811 | 12 | A | C | 53308932 | 0.0789 | 0.1267 | KRT8 | intron_variant |
| rs187809440 | 12 | T | C | 53329231 | 0.2268 | 0.0003 | KRT8 | intron_variant |
| rs7968403 | 12 | T | C | 65012824 | 0.0265 | 0.6390 | RASSF3 | intron_variant;NMD_transcript_variant;intron_variant;NMD_transcript_variant |
| rs4842687 | 12 | A | G | 90156377 | 0.0303 | 0.7122 |  | ; |
| rs1327653 | 13 | T | C | 51076440 | 0.0295 | 0.2546 | DLEU1 | non_coding_transcript_variant;intron_variant |
| rs7489409 | 13 | C | T | 73716861 | 0.0414 | 0.1847 |  |  |
| rs7336001 | 13 | G | C | 73995877 | 0.0554 | 0.9055 | LINC00393 | intron_variant;non_coding_transcript_variant |
| rs1004030 | 14 | T | C | 23305649 | 0.0146 | 0.5830 |  | ; |
| rs6571758 | 14 | G | A | 37136194 | 0.0324 | 0.6195 | PAX9 | intron_variant;non_coding_transcript_variant |
| rs11849126 | 14 | G | A | 38144592 | 0.0026 | 0.6944 | TTC6 | intron_variant;NMD_transcript_variant |
| rs4901313 | 14 | G | T | 53387109 | 0.0401 | 0.8151 | FERMT2 | intron_variant;NMD_transcript_variant |
| rs8005621 | 14 | G | A | 61106699 | 0.0385 | 0.0947 |  |  |
| rs79133931 | 14 | T | C | 64687926 | -0.1985 | 0.0010 | SYNE2 | intron_variant;NMD_transcript_variant;non_coding_transcript_variant;intron_variant |
| rs2093202 | 14 | A | G | 68923908 | 0.0160 | 0.6143 | RAD51B | intron_variant;NMD_transcript_variant;non_coding_transcript_variant |
| rs767127 | 14 | G | A | 69134264 | 0.0238 | 0.4978 | RAD51B | intron_variant;non_coding_transcript_variant |
| rs17565772 | 14 | G | A | 70756333 | 0.0246 | 0.4595 |  | non_coding_transcript_variant;intron_variant |
| rs11561564 | 15 | G | A | 40965044 | 0.0262 | 0.8413 |  |  |
| rs33984059 | 15 | A | G | 56385868 | 0.0808 | 0.9754 | RFX7 | missense_variant;NMD_transcript_variant |
| rs74634457 | 15 | G | A | 66835704 | 0.0380 | 0.2563 | ZWILCH | intron_variant |
| rs12913603 | 15 | A | C | 70668824 | 0.0195 | 0.4757 |  |  |
| rs7188897 | 16 | T | C | 54469331 | 0.0145 | 0.3503 |  |  |
| rs13380763 | 16 | C | T | 54678305 | 0.0265 | 0.8116 |  |  |
| rs11863709 | 16 | C | T | 57654576 | 0.0594 | 0.9647 | GPR56 | intron_variant;non_coding_transcript_variant;NMD_transcript_variant |
| rs28709974 | 16 | C | T | 79847632 | 0.0517 | 0.0528 |  | non_coding_transcript_variant;intron_variant |
| rs8052913 | 16 | C | T | 82166181 | 0.0315 | 0.3854 |  | non_coding_transcript_variant;intron_variant |
| rs72811270 | 17 | A | G | 12585459 | 0.0382 | 0.1155 |  | intron_variant;non_coding_transcript_variant;NMD_transcript_variant;intron_variant |
| rs4795646 | 17 | G | A | 30092898 | 0.0294 | 0.7781 |  | non_coding_transcript_variant;intron_variant |
| rs3110641 | 17 | A | G | 36047417 | 0.0120 | 0.2274 | HNF1B | intron_variant;intron_variant |
| rs11649743 | 17 | G | A | 36074979 | 0.0482 | 0.8100 | HNF1B | intron_variant |
| rs718961 | 17 | A | G | 36077099 | -0.0171 | 0.2301 | HNF1B | intron_variant |
| rs11651052 | 17 | A | G | 36102381 | -0.0153 | 0.4697 | HNF1B | intron_variant |
| rs11263763 | 17 | A | G | 36103565 | 0.0816 | 0.5295 | HNF1B | intron_variant;intron_variant |
| chr17_46820676 | 17 | T | C | 46820676 | 0.0944 | 0.0410 |  |  |
| rs2960158 | 17 | T | C | 47380305 | 0.0114 | 0.7714 | ZNF652 | intron_variant;NMD_transcript_variant |
| rs565189650 | 17 | T | C | 47398245 | 0.0493 | 0.0755 | ZNF652 | intron_variant;NMD_transcript_variant;intron_variant;NMD_transcript_variant |
| rs12938538 | 17 | T | C | 56426027 | 0.0172 | 0.5631 | BZRAP1-AS1 | non_coding_transcript_variant;intron_variant;NMD_transcript_variant;intron_variant |
| rs684232 | 17 | C | T | 618965 | 0.0538 | 0.3546 | VPS53 | intron_variant |
| rs9889335 | 17 | T | G | 69115146 | 0.0288 | 0.4817 | CASC17 | non_coding_transcript_variant;intron_variant |
| rs148511027 | 17 | G | GTTAT | 69117532 | 0.0585 | 0.4745 | CASC17 | non_coding_transcript_variant;intron_variant |
| rs78378222 | 17 | G | T | 7571752 | 0.1025 | 0.0107 | TP53 | intron_variant;non_coding_transcript_exon_variant;3_prime_UTR_variant;3_prime_UTR_variant;intron_variant;non_coding_transcript_exon_variant;3_prime_UTR_variant;3_prime_UTR_variant |
| rs28441558 | 17 | C | T | 7803118 | 0.0623 | 0.0568 | CHD3 | intron_variant |
| rs8089411 | 18 | C | T | 51771322 | 0.0161 | 0.4421 |  |  |
| rs35283980 | 18 | G | C | 56745999 | 0.0131 | 0.3023 |  |  |
| rs533722308 | 18 | CT | C | 60961193 | 0.0244 | 0.3634 | BCL2 | intron_variant;intron_variant |
| rs11876000 | 18 | T | G | 73035513 | 0.0228 | 0.4137 |  |  |
| rs9959454 | 18 | A | G | 76770820 | 0.0489 | 0.7318 |  |  |
| rs10412482 | 19 | C | T | 17228554 | 0.0338 | 0.7162 | MYO9B | intron_variant |
| rs17501397 | 19 | C | T | 32168343 | 0.0217 | 0.9097 |  |  |
| rs59710626 | 19 | G | T | 38548094 | 0.0094 | 0.8637 | SIPA1L3 | intron_variant;non_coding_transcript_variant |
| rs4802297 | 19 | G | C | 38738130 | 0.0368 | 0.4976 | SPINT2 | intron_variant |
| chr19_41985587 | 19 | A | G | 41985587 | -0.0470 | 0.2589 |  | intron_variant;non_coding_transcript_variant |
| rs11673591 | 19 | T | A | 41985931 | 0.0036 | 0.7391 |  | intron_variant;non_coding_transcript_variant |
| rs2659051 | 19 | G | C | 51345568 | 0.0421 | 0.7945 |  | intron_variant;non_coding_transcript_variant |
| rs61752561 | 19 | G | A | 51361382 | 0.1099 | 0.9565 | KLK3 | missense_variant;3_prime_UTR_variant;NMD_transcript_variant;intron_variant;non_coding_transcript_exon_variant;coding_sequence_variant;3_prime_UTR_variant;NMD_transcript_variant;intron_variant;non_coding_transcript_exon_variant |
| chr19_51361757 | 19 | C | T | 51361757 | -0.1912 | 0.0735 | KLK3 | missense_variant;3_prime_UTR_variant;NMD_transcript_variant;non_coding_transcript_exon_variant;coding_sequence_variant;3_prime_UTR_variant;NMD_transcript_variant;non_coding_transcript_exon_variant |
| rs2847344 | 1 | A | G | 10564675 | 0.0167 | 0.6902 | PEX14 | non_coding_transcript_variant;intron_variant |
| rs1811698 | 1 | C | T | 150772613 | 0.0468 | 0.8913 | CTSK | intron_variant |
| rs607518 | 1 | A | G | 150954671 | 0.0242 | 0.2100 | ANXA9 | 5_prime_UTR_variant |
| rs10127983 | 1 | T | C | 153923276 | 0.0390 | 0.3115 | CRTC2 | intron_variant;NMD_transcript_variant;non_coding_transcript_variant |
| rs56103503 | 1 | T | C | 154980351 | 0.0347 | 0.3841 | ZBTB7B | intron_variant;non_coding_transcript_variant |
| rs147847496 | 1 | C | T | 155118588 | 0.0829 | 0.9783 |  |  |
| rs184104770 | 1 | A | C | 155690186 | -0.0083 | 0.0100 | MSTO1 | intron_variant;intron_variant;non_coding_transcript_variant |
| rs80237341 | 1 | C | G | 157119915 | 0.0593 | 0.0127 |  |  |
| rs6660538 | 1 | A | C | 163295678 | 0.0168 | 0.3681 | NUF2 | intron_variant;non_coding_transcript_variant;NMD_transcript_variant |
| rs10803412 | 1 | C | T | 16376831 | 0.0133 | 0.1711 | CLCNKB | intron_variant |
| rs4075646 | 1 | T | A | 167135941 | 0.0405 | 0.0372 |  |  |
| rs507603 | 1 | A | C | 179897070 | 0.0044 | 0.1580 |  | intron_variant;non_coding_transcript_variant |
| rs34295433 | 1 | CTAAG | C | 183032447 | 0.0250 | 0.5325 | LAMC1 | intron_variant |
| rs138638958 | 1 | TTTTG | T | 204030362 | 0.0173 | 0.5364 |  |  |
| rs4245739 | 1 | A | C | 204518842 | 0.0379 | 0.7297 | MDM4 | 3_prime_UTR_variant;non_coding_transcript_exon_variant;intron_variant;3_prime_UTR_variant;non_coding_transcript_exon_variant;intron_variant |
| rs708723 | 1 | C | T | 205739266 | 0.0268 | 0.4366 | RAB7L1 | 3_prime_UTR_variant;NMD_transcript_variant |
| rs544780844 | 1 | T | C | 46251655 | 0.0301 | 0.1424 |  |  |
| rs7542260 | 1 | T | C | 5743196 | -0.0044 | 0.0533 |  |  |
| rs56391074 | 1 | AT | A | 88210715 | 0.0170 | 0.3664 |  |  |
| rs11480453 | 20 | C | CA | 31347512 | 0.0280 | 0.6164 |  |  |
| rs6141551 | 20 | C | T | 34006970 | 0.0111 | 0.6137 |  |  |
| rs73909841 | 20 | T | C | 49548807 | 0.0352 | 0.9278 |  | intron_variant;non_coding_transcript_variant |
| rs6126986 | 20 | C | T | 52464719 | 0.0388 | 0.4825 |  |  |
| rs381331 | 20 | A | G | 62229989 | 0.0178 | 0.6224 | GMEB2 | intron_variant |
| chr20_62233638 | 20 | G | A | 62233638 | -0.0118 | 0.4016 | GMEB2 | intron_variant |
| rs3787099 | 20 | A | G | 62307517 | 0.0800 | 0.9156 | RTEL1 | intron_variant;NMD_transcript_variant;intron_variant |
| rs1058319 | 20 | C | T | 62374389 | 0.0392 | 0.8611 | SLC2A4RG | 3_prime_UTR_variant;non_coding_transcript_exon_variant |
| rs11701433 | 21 | C | T | 40296411 | 0.0187 | 0.3265 |  | intron_variant;non_coding_transcript_variant |
| rs61735792 | 21 | A | G | 42866332 | 0.1249 | 0.0139 | TMPRSS2 | synonymous_variant;intron_variant;non_coding_transcript_variant |
| rs9978557 | 21 | C | T | 42882462 | 0.0600 | 0.9001 | TMPRSS2 | intron_variant |
| rs1978060 | 22 | G | A | 19749525 | 0.0290 | 0.6086 | TBX1 | intron_variant;non_coding_transcript_variant;intron_variant |
| rs9625483 | 22 | A | G | 28888939 | 0.0625 | 0.0247 | TTC28 | intron_variant |
| rs138708 | 22 | G | A | 39138332 | 0.0418 | 0.9795 |  | intron_variant;non_coding_transcript_variant;missense_variant;non_coding_transcript_exon_variant |
| rs34584683 | 22 | T | A | 40499107 | 0.0246 | 0.2073 | TNRC6B | intron_variant |
| rs6003062 | 22 | G | A | 43499741 | 0.0026 | 0.9308 |  |  |
| rs5759167 | 22 | G | T | 43500212 | 0.0631 | 0.5003 |  |  |
| chr22_43501620 | 22 | C | T | 43501620 | -0.0779 | 0.0736 |  |  |
| chr22_43503547 | 22 | C | T | 43503547 | -0.0108 | 0.4232 |  |  |
| rs9615099 | 22 | T | A | 45698149 | 0.0191 | 0.7483 |  |  |
| rs17321482 | 23 | C | T | 11482634 | 0.0249 | 0.8710 | ARHGAP6 | intron_variant;NMD_transcript_variant;non_coding_transcript_variant |
| rs5972255 | 23 | T | C | 30896320 | 0.0085 | 0.2403 | TAB3 | intron_variant |
| rs4907775 | 23 | G | A | 51263200 | 0.0484 | 0.3575 |  |  |
| rs5943724 | 23 | G | A | 52695895 | 0.0081 | 0.6578 |  |  |
| rs4826594 | 23 | A | G | 54454406 | -0.0046 | 0.0586 |  |  |
| chrX_66751555 | 23 | G | A | 66751555 | -0.0288 | 0.1555 |  |  |
| rs5919393 | 23 | T | C | 66825357 | 0.0141 | 0.8457 | AR | intron_variant;non_coding_transcript_variant;NMD_transcript_variant |
| rs11795627 | 23 | T | C | 69957441 | -0.0138 | 0.4819 | TEX11 | intron_variant |
| rs371707439 | 23 | A | G | 70139908 | 0.0217 | 0.1969 |  |  |
| rs960417 | 23 | A | G | 9811095 | 0.0214 | 0.7182 | SHROOM2 | intron_variant |
| rs73913932 | 2 | G | A | 10094526 | 0.0591 | 0.0742 | GRHL1 | NMD_transcript_variant;intron_variant;non_coding_transcript_variant |
| rs1990613 | 2 | T | C | 10781975 | 0.0401 | 0.5153 | NOL10 | intron_variant |
| rs2165108 | 2 | A | T | 111861993 | 0.0385 | 0.0420 |  | intron_variant;non_coding_transcript_variant;intron_variant |
| rs11691517 | 2 | T | G | 111893096 | 0.0336 | 0.7489 | BCL2L11 | intron_variant;NMD_transcript_variant |
| rs111595856 | 2 | T | C | 121103598 | 0.0639 | 0.0676 |  |  |
| rs10206072 | 2 | G | A | 121373466 | 0.0457 | 0.9009 |  |  |
| rs7602028 | 2 | C | A | 16016503 | 0.0135 | 0.7221 |  |  |
| rs16854905 | 2 | C | T | 169012955 | 0.0126 | 0.9021 | STK39 | intron_variant |
| rs77167534 | 2 | C | T | 173319930 | 0.1191 | 0.9411 | ITGA6 | intron_variant |
| rs34925593 | 2 | C | T | 174234547 | 0.0205 | 0.4899 |  |  |
| rs1861270 | 2 | G | A | 202126615 | 0.0202 | 0.7350 | CASP8 | intron_variant;non_coding_transcript_variant;NMD_transcript_variant;intron_variant |
| rs12621900 | 2 | C | T | 208118301 | 0.0236 | 0.7631 |  | non_coding_transcript_variant;intron_variant |
| rs9306894 | 2 | G | A | 20878105 | 0.0148 | 0.3690 |  | non_coding_transcript_exon_variant;non_coding_transcript_exon_variant |
| rs74001374 | 2 | C | T | 238411293 | 0.1019 | 0.9926 | MLPH | intron_variant;non_coding_transcript_variant;intron_variant |
| rs2292884 | 2 | G | A | 238443226 | 0.0354 | 0.2415 | MLPH | intron_variant;missense_variant;non_coding_transcript_variant;non_coding_transcript_exon_variant;missense_variant;intron_variant;coding_sequence_variant;non_coding_transcript_variant;non_coding_transcript_exon_variant;coding_sequence_variant |
| rs77559646 | 2 | A | G | 242135265 | 0.1676 | 0.0227 | ANO7 | missense_variant;splice_region_variant;intron_variant |
| rs77482050 | 2 | G | A | 242139600 | 0.2496 | 0.9892 | ANO7 | missense_variant;stop_gained |
| rs2074840 | 2 | C | T | 242141719 | 0.0210 | 0.3041 | ANO7 | splice_region_variant;synonymous_variant |
| rs76832527 | 2 | A | G | 242157241 | 0.0458 | 0.1740 | ANO7 | missense_variant |
| rs6738169 | 2 | C | G | 43064555 | 0.0286 | 0.7073 |  |  |
| rs7591218 | 2 | A | G | 43637998 | 0.0307 | 0.3130 | THADA | intron_variant;NMD_transcript_variant;non_coding_transcript_variant |
| rs28514770 | 2 | C | G | 43851282 | 0.0139 | 0.7124 |  |  |
| rs11125927 | 2 | G | A | 62752975 | 0.0327 | 0.1139 |  |  |
| rs58235267 | 2 | G | C | 63277843 | 0.0203 | 0.4733 | OTX1 | intron_variant |
| chr2_63301164 | 2 | A | G | 63301164 | -0.0327 | 0.4982 |  |  |
| rs139283528 | 2 | G | A | 63938756 | 0.1070 | 0.9882 | WDPCP | non_coding_transcript_variant;intron_variant |
| rs74702681 | 2 | T | C | 66652885 | 0.0564 | 0.0225 |  |  |
| rs2028900 | 2 | C | T | 85767735 | 0.0384 | 0.5539 | MAT2A | intron_variant;non_coding_transcript_variant |
| rs11686272 | 2 | T | G | 8598444 | 0.0180 | 0.4473 |  |  |
| rs1283104 | 3 | G | C | 106962521 | 0.0154 | 0.3755 | LINC00883 | intron_variant;non_coding_transcript_variant |
| rs151038334 | 3 | C | T | 107193337 | 0.0431 | 0.9157 |  |  |
| rs2271494 | 3 | A | T | 113300183 | 0.0474 | 0.5699 | SIDT1 | intron_variant;non_coding_transcript_variant |
| rs2811476 | 3 | C | A | 127898501 | 0.0096 | 0.2660 | EEFSEC | intron_variant;non_coding_transcript_variant |
| rs4857841 | 3 | A | G | 128046643 | 0.0427 | 0.2779 | EEFSEC | intron_variant;non_coding_transcript_variant |
| rs35006112 | 3 | G | A | 128213994 | 0.0296 | 0.8415 |  | intron_variant;non_coding_transcript_variant |
| rs1457063 | 3 | A | G | 137562823 | 0.0182 | 0.6146 |  |  |
| rs7650602 | 3 | C | T | 141147414 | 0.0135 | 0.4413 | ZBTB38 | intron_variant |
| rs2293607 | 3 | T | C | 169482335 | 0.0284 | 0.7525 | TERC | non_coding_transcript_exon_variant;non_coding_transcript_exon_variant |
| rs78416326 | 3 | G | C | 170074517 | 0.1039 | 0.7929 |  | ; |
| rs577952184 | 3 | C | CTTTTT | 170083540 | 0.0331 | 0.8809 | SKIL | intron_variant;NMD_transcript_variant |
| rs6550597 | 3 | A | G | 18738940 | 0.0135 | 0.7138 |  | non_coding_transcript_variant;intron_variant |
| rs7618603 | 3 | A | C | 23153062 | 0.0205 | 0.1704 |  |  |
| rs34680713 | 3 | A | AT | 49621718 | 0.0300 | 0.1756 | BSN | intron_variant |
| rs13091518 | 3 | T | C | 70796696 | 0.0264 | 0.5766 |  |  |
| rs143745027 | 3 | G | A | 87144017 | 0.0597 | 0.0650 |  |  |
| chr3_87147922 | 3 | T | A | 87147922 | -0.0020 | 0.9268 |  |  |
| rs7628934 | 3 | C | T | 87175984 | 0.0189 | 0.5055 |  |  |
| rs6788616 | 3 | G | A | 87205079 | 0.0301 | 0.4599 |  |  |
| rs114810266 | 3 | A | G | 87399362 | 0.0438 | 0.9775 |  |  |
| rs7679673 | 4 | C | A | 106061534 | 0.0668 | 0.5983 |  | non_coding_transcript_variant;intron_variant |
| rs17035310 | 4 | C | T | 106064754 | 0.0243 | 0.8683 |  |  |
| rs77821238 | 4 | C | T | 140948835 | 0.0156 | 0.8315 | MAML3 | intron_variant |
| rs72725734 | 4 | G | A | 146879237 | 0.0102 | 0.1402 |  |  |
| rs147762399 | 4 | T | C | 152030340 | 0.0429 | 0.0363 | SH3D19 | non_coding_transcript_variant;intron_variant |
| rs17804499 | 4 | G | C | 74442349 | 0.0502 | 0.9437 | RASSF6 | missense_variant |
| rs13142786 | 4 | T | A | 74477135 | 0.0275 | 0.4797 | RASSF6 | intron_variant;non_coding_transcript_variant |
| rs6853490 | 4 | G | A | 95544718 | 0.0218 | 0.4364 | PDLIM5 | intron_variant;non_coding_transcript_variant;NMD_transcript_variant |
| chr4_95562877 | 4 | T | C | 95562877 | -0.0317 | 0.3507 | PDLIM5 | intron_variant;non_coding_transcript_variant;NMD_transcript_variant |
| rs2242652 | 5 | G | A | 1280028 | 0.0198 | 0.8007 | TERT | non_coding_transcript_variant;intron_variant;NMD_transcript_variant;intron_variant;non_coding_transcript_variant;intron_variant;NMD_transcript_variant;intron_variant |
| rs7725218 | 5 | A | G | 1282414 | -0.0393 | 0.3501 | TERT | non_coding_transcript_variant;intron_variant;NMD_transcript_variant;intron_variant |
| rs71595003 | 5 | A | G | 1292118 | 0.0815 | 0.0285 | TERT | intron_variant;NMD_transcript_variant;intron_variant |
| rs2736098 | 5 | T | C | 1294086 | 0.0348 | 0.2626 | TERT | synonymous_variant;NMD_transcript_variant;synonymous_variant |
| rs2736108 | 5 | T | C | 1297488 | 0.0173 | 0.2944 |  | ; |
| rs10793821 | 5 | T | C | 133836209 | 0.0195 | 0.5760 |  |  |
| rs71599622 | 5 | T | TGA | 14372362 | 0.0136 | 0.2826 | TRIO | intron_variant;NMD_transcript_variant;non_coding_transcript_variant |
| rs76551843 | 5 | A | G | 169172133 | 0.1520 | 0.9913 | DOCK2 | intron_variant;NMD_transcript_variant;non_coding_transcript_variant |
| rs9686557 | 5 | C | A | 172959030 | 0.0216 | 0.4429 |  |  |
| rs61739424 | 5 | G | A | 177683905 | 0.0495 | 0.9624 | COL23A1 | missense_variant |
| rs2672843 | 5 | G | A | 177891551 | 0.0207 | 0.4090 | COL23A1 | intron_variant |
| rs4975758 | 5 | G | C | 1891174 | 0.0254 | 0.4757 |  | non_coding_transcript_variant;intron_variant |
| rs10941370 | 5 | T | C | 37833419 | 0.0081 | 0.4452 | GDNF | intron_variant |
| rs1482675 | 5 | T | C | 44368506 | 0.0054 | 0.3113 | FGF10 | intron_variant |
| rs9292122 | 5 | A | G | 56087910 | 0.0181 | 0.7106 |  |  |
| rs2038542 | 6 | C | T | 109295293 | 0.0352 | 0.1455 |  |  |
| rs2018336 | 6 | T | C | 11217897 | 0.0415 | 0.7696 |  | intron_variant;non_coding_transcript_variant;intron_variant;NMD_transcript_variant |
| rs339351 | 6 | C | A | 117200434 | 0.0409 | 0.6990 | RFX6 | non_coding_transcript_variant;intron_variant |
| rs3910736 | 6 | T | C | 153412476 | -0.0212 | 0.3242 | RGS17 | intron_variant |
| rs13215045 | 6 | C | T | 153447516 | 0.0202 | 0.6868 | RGS17 | intron_variant |
| rs963800 | 6 | C | T | 160150279 | 0.0285 | 0.7868 | SOD2 | intron_variant;intron_variant;non_coding_transcript_variant |
| rs4646284 | 6 | TG | T | 160581543 | 0.0942 | 0.2974 |  |  |
| rs7769879 | 6 | C | G | 160865645 | 0.0325 | 0.3645 | SLC22A3 | intron_variant |
| rs2814811 | 6 | A | G | 1670985 | 0.0187 | 0.4043 | GMDS | intron_variant;non_coding_transcript_variant |
| rs6927369 | 6 | C | T | 21330689 | 0.0333 | 0.8084 |  |  |
| rs4269363 | 6 | G | A | 21471490 | 0.0167 | 0.5721 |  |  |
| rs12665509 | 6 | A | C | 21878849 | 0.0150 | 0.4603 | CASC15 | intron_variant;non_coding_transcript_variant |
| rs62407547 | 6 | C | T | 30216712 | 0.0212 | 0.2467 | HCG17 | intron_variant;non_coding_transcript_variant |
| rs9275160 | 6 | A | G | 32652620 | 0.0371 | 0.3525 |  |  |
| rs9469899 | 6 | A | G | 34793124 | 0.0333 | 0.3648 | UHRF1BP1 | intron_variant |
| rs4714485 | 6 | G | T | 41536587 | 0.0384 | 0.2754 | FOXP4 | intron_variant |
| rs9472120 | 6 | C | T | 43709785 | 0.0173 | 0.4902 |  |  |
| rs9443189 | 6 | A | G | 76495882 | 0.0116 | 0.8596 | MYO6 | intron_variant |
| rs4513875 | 7 | T | C | 1928159 | 0.0240 | 0.3860 | MAD1L1 | intron_variant |
| rs11452686 | 7 | T | TA | 20414110 | 0.0005 | 0.5922 | ITGB8 | intron_variant;non_coding_transcript_variant |
| rs9655205 | 7 | C | A | 20999211 | 0.0448 | 0.2274 | LINC01162 | intron_variant;non_coding_transcript_variant |
| rs35389879 | 7 | T | G | 21812043 | 0.0238 | 0.4149 | DNAH11 | intron_variant |
| rs6956484 | 7 | A | C | 27564862 | 0.0335 | 0.6145 |  |  |
| rs10486567 | 7 | G | A | 27976563 | 0.0460 | 0.7708 | JAZF1 | intron_variant;NMD_transcript_variant |
| rs12701838 | 7 | A | G | 40877473 | 0.0275 | 0.7309 | SUGCT | intron_variant;non_coding_transcript_variant |
| rs834608 | 7 | A | T | 47451918 | 0.0243 | 0.5760 | TNS3 | intron_variant |
| rs6955627 | 7 | C | T | 92577760 | 0.0216 | 0.9154 |  |  |
| rs4727386 | 7 | A | G | 97688440 | 0.0188 | 0.4545 |  | ; |
| rs6965016 | 7 | C | A | 97807882 | 0.0337 | 0.4591 | LMTK2 | intron_variant |
| rs73700335 | 8 | G | A | 108923107 | 0.0308 | 0.8343 | RSPO2 | intron_variant |
| rs2572375 | 8 | C | T | 11217455 | 0.0152 | 0.7046 | TDH | non_coding_transcript_variant;intron_variant |
| rs6984837 | 8 | G | A | 127901649 | 0.0026 | 0.3184 | PCAT1 | intron_variant;non_coding_transcript_variant |
| rs9297746 | 8 | C | T | 127909361 | -0.0391 | 0.5029 | PCAT1 | intron_variant;non_coding_transcript_variant |
| rs7011138 | 8 | A | T | 127922200 | -0.0162 | 0.8196 | PCAT1 | intron_variant;non_coding_transcript_variant |
| rs28556804 | 8 | G | A | 128014315 | -0.0592 | 0.2627 | PCAT1 | intron_variant;non_coding_transcript_variant |
| rs7463326 | 8 | G | A | 128027954 | 0.0069 | 0.7470 | PCAT1 | intron_variant;non_coding_transcript_variant |
| rs77541621 | 8 | A | G | 128077146 | 0.2435 | 0.0251 |  |  |
| rs1016343 | 8 | T | C | 128093297 | -0.0015 | 0.2058 | PCAT2 | non_coding_transcript_variant;intron_variant |
| rs72725879 | 8 | T | C | 128103969 | 0.0994 | 0.1849 |  |  |
| rs183373024 | 8 | G | A | 128104117 | 0.3390 | 0.0063 |  |  |
| rs78809737 | 8 | G | A | 128104218 | 0.0370 | 0.9540 |  |  |
| rs60163266 | 8 | A | G | 128323157 | 0.0184 | 0.1382 | CASC8 | intron_variant;non_coding_transcript_variant |
| rs201057014 | 8 | T | C | 128325355 | 0.0110 | 0.1109 | CASC8 | intron_variant;non_coding_transcript_variant |
| rs17464492 | 8 | A | G | 128342866 | 0.0642 | 0.7142 | CASC8 | intron_variant;non_coding_transcript_variant |
| rs6983267 | 8 | G | T | 128413305 | 0.0832 | 0.5138 | CASC8 | intron_variant;non_coding_transcript_variant;intron_variant;non_coding_transcript_variant |
| rs7812894 | 8 | T | A | 128520479 | -0.0862 | 0.8998 |  |  |
| rs10090154 | 8 | T | C | 128532137 | 0.0492 | 0.0997 |  |  |
| rs34265760 | 8 | T | C | 128535543 | 0.0466 | 0.9806 |  |  |
| rs12549761 | 8 | C | G | 128540776 | 0.0899 | 0.8769 |  |  |
| rs6557704 | 8 | A | G | 23470785 | 0.0285 | 0.2839 |  |  |
| chr8_23525358 | 8 | C | T | 23525358 | 0.0519 | 0.4314 |  |  |
| rs12677206 | 8 | A | C | 26063165 | 0.0152 | 0.3396 |  |  |
| rs11467 | 8 | A | G | 38644914 | 0.0178 | 0.6294 | TACC1 | intron_variant;non_coding_transcript_variant;5_prime_UTR_variant |
| rs870167 | 8 | G | A | 8498803 | 0.0256 | 0.0648 |  |  |
| rs4451364 | 9 | A | G | 109532734 | 0.0281 | 0.7642 |  |  |
| rs817872 | 9 | C | T | 110144887 | 0.0298 | 0.2772 |  |  |
| rs143655302 | 9 | G | A | 110290217 | 0.1524 | 0.9855 |  |  |
| rs2241167 | 9 | A | G | 130430116 | 0.0244 | 0.5134 | STXBP1 | intron_variant |
| rs12634 | 9 | T | G | 132573536 | 0.0342 | 0.2361 | TOR1B | 3_prime_UTR_variant |
| rs34540271 | 9 | C | T | 18554773 | 0.0132 | 0.6956 | ADAMTSL1 | intron_variant |
| rs10122990 | 9 | C | A | 19072246 | 0.0190 | 0.3753 | HAUS6 | intron_variant |
| rs17694493 | 9 | G | C | 22041998 | 0.0260 | 0.1366 | CDKN2B-AS1 | non_coding_transcript_variant;intron_variant |
| rs10122495 | 9 | T | A | 34049779 | 0.0032 | 0.2858 |  |  |
| rs139135938 | 20 | TGGCAGTGGCAGC | T | 61001061 | 0.0250 | 0.6818 | RBBP8NL | intron_variant |
| rs555607708 | 22 | A | AG | 29091856 | 0.1694 | 0.0024 | CHEK2 | frameshift_variant;intron_variant;3_prime_UTR_variant;NMD_transcript_variant |
| rs145053401 | 3 | C | CAT | 152011745 | 0.0435 | 0.8924 | MBNL1 | non_coding_transcript_variant;intron_variant |
| rs150184171 | 6 | G | GTGTTGT | 134292717 | 0.0231 | 0.5643 | TBPL1 | intron_variant |
| rs57588856 | 8 | C | CAA | 127836925 | 0.0221 | 0.6435 |  |  |
| rs142727307 | 9 | T | TG | 82090723 | 0.0178 | 0.1115 |  |  |
| rs11338635 | 23 | GA | G | 51245276 | 0.0032 | 0.3563 |  | intron_variant;non_coding_transcript_variant |
| rs141811748 | 8 | C | CCAAA | 25894201 | 0.0487 | 0.1442 |  |  |

**Additional PRACTICAL Consortium Collaborators**

Teuvo L J Tammela^1,2^

Anssi Auvinen^3^

Alison M Dunning^4^

Maya Ghoussaini^5^

Suzanne Chambers^6,7^

Lisa Horvath^8,9^

Leire Moya^10,11^

Gail P Risbridger^12,13^

Wayne Tilley^14^

Judith A Clements^10,11^

Markus Aly^15,16,17^

Tobias Nordström^18,19^

Jenny L Donovan^20^

Freddie C Hamdy^21,22^

Richard M Martin^20,23,24^

Stig E Bojesen^25,26^

Peter Iversen^27^

Martin Andreas Røder^27^

Ian M Thompson Jr^28^

Melissa C Southey^29,30,31^

Graham G Giles^30,32,29^

Roger L Milne^30,32,29^

Niclas Håkansson^33^

Stephanie Weinstein^34^

Fredrick R Schumacher^35,36^

Loic Le Marchand^37^

Xin Sheng^38^

Tim J Key^39^

Elaine A Ostrander^40^

Phyllis J Goodman^41^

Edward Giovannucci^42^

Neil Burnet^43^

Gill Barnett^44^

Bettina F Drake^45^

Géraldine Cancel-Tassin^46,47^

Stephen Chanock^34^

Gerald L Andriole^48^

Robert N Hoover^34^

Mitchell J Machiela^34^

Laura E Beane Freeman^34^

Michael Borre^49,50^

Dominika Woko?orczyk^51^

Jan Lubinski^51^

Lovise Maehle^52^

Thérèse Truong^53^

Hui-Yi Lin^54^

Mariana C Stern^55^

Manuel Luedeke^56^

Thomas Schnoeller^57^

Neil E Fleshner^58^

Antonio Finelli^59^

Sarah L Kerns^60^

Harry Ostrer^61^

Hong-Wei Zhang^62^

Ninghan Feng^63^

Xin Guo^64^

Xueying Mao^65^

Zan Sun^66^

Guoping Ren^67^

Antonio Gómez-Caamaño^68^

Laura Fachal^69,70,71^

Jeannette T Bensen^72,73^

Elizabeth TH Fontham^74^

James L Mohler^75,73^

Jack A Taylor^76,77^

Javier Llorca^78,79^

Gemma Castaño-Vinyals^80,81,82^

Antonio Alcaraz^83^

Robert Szulkin^84,85^

Martin Eklund^86^

Peter Kraft^87^

Sara Lindström^88^

Meir Stampfer^89^

Paula Paulo^90^

Andreia Brandão^91,90^

Marta Cardoso^90^

Maria P Silva^91,90^

Pascal Blanchet^92^

Laurent Brureau^92^

Xin Gao^93^

Bernd Holleczek^94^

Ben Schöttker^93^

Chavdar Slavov^95^

Vanio Mitev^96^

Jeri Kim^97^

Yuan Chun Ding^98^

Linda Steele^98^

Gert De Meerleer^99^

Jasmine Lim^100^

Soo-Hwang Teo^101^

Daniel W Lin^102,103^

Davor Lessel^104^

Tomislav Kulis^105^

Nawaid Usmani^106,107^

Matthew Parliament^106,107^

Steven Joniau^108^

Thomas Van den Broeck^109^

Manuela Gago-Dominguez^110,108^

Maria Elena Martinez^111,111^

William S Bush^112^

Melinda C Aldrich^113^

Jennifer Cullen^35^

Monique J Roobol^114^

Guido Jenster^114^

Maureen Sanderson^115^

^1^Department of Urology, Tampere University Hospital, FI-33521 Tampere, Finland

^2^Faculty of Medicine and Health Technology, Tampere University, FI-33100 Tampere, Finland

^3^Unit of Health Sciences, Faculty of Social Sciences, Tampere University, Tampere, Finland

^4^Centre for Cancer Genetic Epidemiology, Department of Oncology, University of Cambridge, Strangeways Laboratory, Worts Causeway, Cambridge, CB1 8RN, UK

^5^Open Targets, Wellcome Sanger Institute, Hinxton, Saffron Walden, CB10 1SA, UK

^6^University of Technology, Sydney

^7^Cancer Council Queensland, Fortitude Valley, QLD 4006, Australia

^8^Chris O'Brien Lifehouse (COBLH), Camperdown, Sydney, NSW 2010, Australia

^9^Garvan Institute of Medical Research, Sydney NSW 2010, Australia

^10^Australian Prostate Cancer Research Centre-Qld, Institute of Health and Biomedical Innovation and School of Biomedical Sciences, Queensland University of Technology, Brisbane QLD 4059, Australia

^11^Translational Research Institute, Brisbane, Queensland 4102, Australia

^12^Department of Anatomy and Developmental Biology, Biomedicine Discovery Institute, Monash University, Melbourne,Victoria 3800, Australia

^13^Prostate Cancer Translational Research Program, Cancer Research Division, Peter MacCallum Cancer Centre, Melbourne, VIC 3000, Australia

^14^Dame Roma Mitchell Cancer Research Laboratories, University of Adelaide, Adelaide, South Australia, Australia

^15^Department of Medical Epidemiology and Biostatistics, Karolinska Institute, SE-171 77 Stockholm, Sweden

^16^Department of Molecular Medicine and Surgery, Karolinska Institutet

^17^Department of Urology, Karolinska University Hospital, Solna, 171 76 Stockholm

^18^Department of Medical Epidemiology and Biostatistics, Karolinska Institute, Stockholm, Sweden

^19^Department of Clinical Sciences at Danderyds Hospital, Karolinska Institutet, Stockholm, Sweden

^20^Population Health Sciences, Bristol Medical School, University of Bristol, BS8 2PS, UK

^21^Nuffield Department of Surgical Sciences, University of Oxford, Oxford, OX1 2JD, UK

^22^Faculty of Medical Science, University of Oxford, John Radcliffe Hospital, Oxford, UK

^23^National Institute for Health Research (NIHR) Biomedical Research Centre, University of Bristol, Bristol, BS8 1TH, UK

^24^Medical Research Council (MRC) Integrative Epidemiology Unit, University of Bristol, Bristol, BS8 2BN, UK.

^25^Faculty of Health and Medical Sciences, University of Copenhagen, 2200 Copenhagen, Denmark

^26^Department of Clinical Biochemistry, Herlev and Gentofte Hospital, Copenhagen University Hospital, Herlev, 2200 Copenhagen, Denmark

^27^Copenhagen Prostate Cancer Center, Department of Urology, Rigshospitalet, Copenhagen University Hospital, DK-2730 Herlev, Copenhagen, Denmark

^28^CHRISTUS Santa Rosa Hospital – Medical Center, San Antonio, TX, USA

^29^Precision Medicine, School of Clinical Sciences at Monash Health, Monash University, Clayton, Victoria 3168, Australia

^30^Cancer Epidemiology Division, Cancer Council Victoria, 615 St Kilda Road, Melbourne, VIC 3004, Australia

^31^Department of Clinical Pathology, The Melbourne Medical School, The University of Melbourne, Melbourne, VIC 3010, Australia.

^32^Centre for Epidemiology and Biostatistics, Melbourne School of Population and Global Health, The University of Melbourne, Grattan Street, Parkville, VIC 3010, Australia

^33^Unit of Cardiovascular and Nutritional Epidemiology, Institute of Environmental Medicine, Karolinska Institutet, SE-171 77 Stockholm, Sweden

^34^Division of Cancer Epidemiology and Genetics, National Cancer Institute, NIH, Bethesda, Maryland, 20892, USA

^35^Department of Population and Quantitative Health Sciences, Case Western Reserve University, Cleveland, OH 44106-7219, USA

^36^Seidman Cancer Center, University Hospitals, Cleveland, OH 44106, USA.

^37^Epidemiology Program, University of Hawaii Cancer Center, Honolulu, HI 96813, USA

^38^Center for Genetic Epidemiology, Department of Preventive Medicine, Keck School of Medicine, University of Southern California/Norris Comprehensive Cancer Center, Los Angeles, CA 90015, USA

^39^Cancer Epidemiology Unit, Nuffield Department of Population Health, University of Oxford, Oxford, OX3 7LF, UK

^40^National Human Genome Research Institute, National Institutes of Health, 50 South Drive, Rm. 5351, Bethesda, MD 20892, USA

^41^SWOG Statistical Center, Fred Hutchinson Cancer Research Center, Seattle, WA 98109, USA

^42^Department of Epidemiology, Harvard School of Public Health, Boston, MA 02115, USA

^43^Division of Cancer Sciences, University of Manchester, Manchester Cancer Research Centre, Manchester Academic Health Science Centre, and The Christie NHS Foundation Trust, Manchester, UK

^44^University of Cambridge, Department of Oncology, Oncology Centre, Cambridge University Hospitals NHS Foundation Trust, Hills Road, Cambridge, CB2 0QQ, UK

^45^Washington University School of Medicine, 660 S. Euclid Avenue, Campus Box 8242, St. Louis, MO 63110 , USA

^46^CeRePP, Tenon Hospital, F-75020 Paris, France.

^47^Sorbonne Universite, GRC n°5 , AP-HP, Tenon Hospital, 4 rue de la Chine, F-75020 Paris, France

^48^The Washington University School of Medicine, 660 S. Euclid Avenue, Campus Box 8242, St. Louis, MO 63110 , USA

^49^Department of Urology, Aarhus University Hospital, Palle Juul-Jensen Boulevard 99, 8200 Aarhus N, Denmark

^50^Department of Clinical Medicine, Aarhus University, DK-8200 Aarhus N

^51^International Hereditary Cancer Center, Department of Genetics and Pathology, Pomeranian Medical University, 70-115 Szczecin, Poland

^52^Department of Medical Genetics, Oslo University Hospital, 0424 Oslo, Norway

^53^Exposome and Heredity, CESP (UMR 1018), Faculté de Médecine, Université Paris-Saclay, Inserm, Gustave Roussy, Villejuif

^54^School of Public Health, Louisiana State University Health Sciences Center, New Orleans, LA 70112, USA

^55^Department of Preventive Medicine, Keck School of Medicine, University of Southern California/Norris Comprehensive Cancer Center, Los Angeles, CA 90015, USA

^56^genetikum, Wegenerstr. 15, D-89231 Neu-Ulm, Germany

^57^Department of Urology, University Hospital Ulm, Germany

^58^Dept. of Surgical Oncology, UHN Princess Margaret Cancer Centre, Toronto ON M5G 2M9, Canada

^59^Division of Urology, Princess Margaret Cancer Centre, Toronto ON M5G 2M9, Canada

^60^Department of Radiation Oncology, University of Rochester Medical Center, Saunders Research Building, Rm 2.202, 265 Crittenden Boulevard, Rochester, NY 14620, USA

^61^Department of Pathology, Albert Einstein College of Medicine, 1300 Morris Park Avenue, Ullman 817, Bronx, NY 10461, USA

^62^Second Military Medical University, 800 Xiangyin Rd., Shanghai 200433, P. R. China

^63^Wuxi Second Hospital,  Nanjing Medical University, 68 Zhongshan Road, Wuxi, Jiangzhu Province, 214003, China

^64^1 Youyi Road, Department of Urology, The First Affiliated Hospital, Chongqing Medical University, Chongqing, 400016, China

^65^Centre for Molecular Oncology, Barts Cancer Institute, Queen Mary University of London, John Vane Science Centre, Charterhouse Square, London EC1M 6BQ, UK

^66^33 Wenyi Road, The People’s Hospital of Liaoning Province, The People’s Hospital of China Medical University, Shenyang, 110016, China

^67^79 Qingchun Road, Department of Pathology, The First Affiliated Hospital, Zhejiang University Medical College, Hangzhou, 310009, China

^68^Department of Radiation Oncology, Complexo Hospitalario Universitario de Santiago, SERGAS, Santiago de Compostela 15706, Spain

^69^Centre for Cancer Genetic Epidemiology, Department of Public Health and Primary Care, University of Cambridge, Strangeways Research Laboratory, Cambridge, CB2 0SR, UK.

^70^Fundación Pública Galega Medicina Xenómica, Santiago De Compostela, 15706, Spain.

^71^Instituto de Investigación Sanitaria de Santiago de Compostela, Santiago De Compostela, 15706, Spain.

^72^Department of Epidemiology, University of North Carolina at Chapel Hill, Chapel Hill, NC, USA

^73^Lineberger Comprehensive Cancer Center, University of North Carolina at Chapel Hill, Chapel Hill, NC, USA

^74^School of Public Health, Louisiana State University Health Sciences Center, New Orleans, LA, USA

^75^Department of Urology, Roswell Park Comprehensive Cancer Center, Buffalo, NY, USA

^76^Epidemiology Branch, National Institute of Environmental Health Sciences, Research Triangle Park, NC, USA

^77^Epigenetic and Stem Cell Biology Laboratory, National Institute of Environmental Health Sciences, Research Triangle Park, NC

^78^University of Cantabria, 39005 Santander, Spain

^79^CIBER Epidemiología y Salud Pública (CIBERESP), 28029 Madrid, Spain

^80^ISGlobal, Barcelona, Spain

^81^IMIM (Hospital del Mar Medical Research Institute), Barcelona, Spain

^82^Universitat Pompeu Fabra (UPF), Barcelona, Spain

^83^Department and Laboratory of Urology. Hospital Clínic. Institut d’Investigacions Biomèdiques August Pi i Sunyer (IDIBAPS), Universitat de Barcelona. Spain. C/Villarroel 170; 08036 Barcelona, Spain

^84^Department of Medical Epidemiology and Biostatistics, Karolinska Institute, Stockholm

^85^SDS Life Science, Danderyd, Sweden

^86^Department of Medical Epidemiology and Biostatistics, Karolinska Institute, 171 77 Stockholm, Sweden

^87^Program in Genetic Epidemiology and Statistical Genetics, Department of Epidemiology, Harvard School of Public Health, Boston, MA, USA

^88^Department of Epidemiology, Health Sciences Building, University of Washington

^89^Channing Division of Network Medicine, Department of Medicine, Brigham and Women's Hospital/Harvard Medical School, Boston, MA 02184, USA

^90^Cancer Genetics Group, IPO-Porto Research Center (CI-IPOP), Portuguese Oncology Institute of Porto (IPO-Porto), Porto, Portugal

^91^Department of Genetics, Portuguese Oncology Institute of Porto (IPO-Porto), Porto, Portugal

^92^CHU de Pointe-à-Pitre, Univ Antilles, Univ Rennes, Inserm, EHESP, Irset (Institut de recherche en santé, environnement et travail) - UMR_S 1085, Pointe-à-Pitre, France

^93^Division of Clinical Epidemiology and Aging Research, German Cancer Research Center (DKFZ), D-69120, Heidelberg, Germany

^94^Saarland Cancer Registry, 66119 Saarbrücken, Germany

^95^Department of Urology and Alexandrovska University Hospital, Medical University of Sofia, 1431 Sofia, Bulgaria

^96^Molecular Medicine Center, Department of Medical Chemistry and Biochemistry, Medical University of Sofia, Sofia, 2 Zdrave Str., 1431 Sofia, Bulgaria

^97^The University of Texas M. D. Anderson Cancer Center, Department of Genitourinary Medical Oncology, 1515 Holcombe Blvd., Houston, TX 77030, USA

^98^Department of Population Sciences, Beckman Research Institute of the City of Hope, 1500 East Duarte Road, Duarte, CA 91010, 626-256-HOPE (4673)

^99^Ghent University, B-9000, Gent

^100^Department of Surgery, Faculty of Medicine, University of Malaya, 50603 Kuala Lumpur, Malaysia

^101^Cancer Research Malaysia (CRM), Outpatient Centre, Subang Jaya Medical Centre, Subang Jaya, Selangor, Malaysia

^102^Division of Public Health Sciences, Fred Hutchinson Cancer Research Center, Seattle, Washington, 98109-1024, USA

^103^Department of Urology, University of Washington, 1959 NE Pacific Street, Box 356510, Seattle, WA 98195, USA

^104^Institute of Human Genetics, University Medical Center Hamburg-Eppendorf, D-20246 Hamburg, Germany

^105^Department of Urology, University Hospital Center Zagreb, University of Zagreb School of Medicine , Zagreb, Croatia

^106^Department of Oncology, Cross Cancer Institute, University of Alberta, 11560 University Avenue, Edmonton, Alberta, Canada T6G 1Z2

^107^Division of Radiation Oncology, Cross Cancer Institute, 11560 University Avenue, Edmonton, Alberta, Canada T6G 1Z2

^108^Department of Urology, University Hospitals Leuven, Herestraat 49, Box 7003 41, BE-3000 Leuven, Belgium

^109^Molecular Endocrinology Laboratory, Department of Cellular and Molecular Medicine, KU Leuven, BE-3000, Belgium

^110^Genomic Medicine Group, Galician Foundation of Genomic Medicine, Instituto de Investigacion Sanitaria de Santiago de Compostela (IDIS), Complejo Hospitalario Universitario de Santiago, Servicio Galego de Saúde, SERGAS, 15706, Santiago de Compostela, Spai

^111^University of California San Diego, Moores Cancer Center, Department of Family Medicine and Public Health, University of California San Diego, La Jolla, CA 92093-0012, USA

^112^Case Western Reserve University, Department of Population and Quantitative Health Sciences, Cleveland Institute for Computational Biology, 2103 Cornell Road, Wolstein Research Building, Suite 2530, Cleveland, OH, 44106 USA

^113^Vanderbilt University Medical Center, Department of Thoracic Surgery, 609 Oxford House, 1313 21st Avenue South, Nashville, TN 37232-4682 USA

^114^Department of Urology, Erasmus University Medical Center, 3015 CE Rotterdam, The Netherlands

^115^Department of Family and Community Medicine, Meharry Medical College, 1005 Dr. DB Todd Jr. Blvd., Nashville, TN 37208 USA

**Funding & Acknowledgements**

CRUK and PRACTICAL consortium

This work was supported by the Canadian Institutes of Health Research, European Commission's Seventh Framework Programme grant agreement n° 223175 (HEALTH-F2-2009-223175), Cancer Research UK Grants C5047/A7357, C1287/A10118, C1287/A16563, C5047/A3354, C5047/A10692, C16913/A6135, and The National Institute of Health (NIH) Cancer Post-Cancer GWAS initiative grant: No. 1 U19 CA 148537-01 (the GAME-ON initiative).

We would also like to thank the following for funding support: The Institute of Cancer Research and The Everyman Campaign, The Prostate Cancer Research Foundation, Prostate Research Campaign UK (now Prostate Action), The Orchid Cancer Appeal, The National Cancer Research Network UK, The National Cancer Research Institute (NCRI) UK. We are grateful for support of NIHR funding to the NIHR Biomedical Research Centre at The Institute of Cancer Research and The Royal Marsden NHS Foundation Trust.

The Prostate Cancer Program of Cancer Council Victoria also acknowledge grant support from The National Health and Medical Research Council, Australia (126402, 209057, 251533, , 396414, 450104, 504700, 504702, 504715, 623204, 940394, 614296,), VicHealth, Cancer Council Victoria, The Prostate Cancer Foundation of Australia, The Whitten Foundation, PricewaterhouseCoopers, and Tattersall’s. EAO, DMK, and EMK acknowledge the Intramural Program of the National Human Genome Research Institute for their support.

Genotyping of the OncoArray was funded by the US National Institutes of Health (NIH) [U19 CA 148537 for ELucidating Loci Involved in Prostate cancer SuscEptibility (ELLIPSE) project and X01HG007492 to the Center for Inherited Disease Research (CIDR) under contract number HHSN268201200008I].

This study would not have been possible without the contributions of the following: Coordination team, bioinformatician and genotyping centers: Genotyping at CCGE, Cambridge: Caroline Baines and Don Conroy

Funding for the iCOGS infrastructure came from: the European Community's Seventh Framework Programme under grant agreement n° 223175 (HEALTH-F2-2009-223175) (COGS), Cancer Research UK (C1287/A10118, C1287/A 10710, C12292/A11174, C1281/A12014, C5047/A8384, C5047/A15007, C5047/A10692, C8197/A16565), the National Institutes of Health (CA128978) and Post-Cancer GWAS initiative (1U19 CA148537, 1U19 CA148065 and 1U19 CA148112 - the GAME-ON initiative), the Department of Defence (W81XWH-10-1-0341), the Canadian Institutes of Health Research (CIHR) for the CIHR Team in Familial Risks of Breast Cancer, Komen Foundation for the Cure, the Breast Cancer Research Foundation, and the Ovarian Cancer Research Fund.

This study would not have been possible without the contributions of the following: Per Hall (COGS); Douglas F. Easton, Paul Pharoah, Kyriaki Michailidou, Manjeet K. Bolla, Qin Wang (BCAC), Andrew Berchuck (OCAC), Rosalind A. Eeles, Douglas F. Easton, Ali Amin Al Olama, Zsofia Kote-Jarai, Sara Benlloch (PRACTICAL), Georgia Chenevix-Trench, Antonis Antoniou, Lesley McGuffog, Fergus Couch and Ken Offit (CIMBA), Joe Dennis, Alison M. Dunning, Andrew Lee, and Ed Dicks, Craig Luccarini and the staff of the Centre for Genetic Epidemiology Laboratory, Javier Benitez, Anna Gonzalez-Neira and the staff of the CNIO genotyping unit, Jacques Simard and Daniel C. Tessier, Francois Bacot, Daniel Vincent, Sylvie LaBoissière and Frederic Robidoux and the staff of the McGill University and Génome Québec Innovation Centre, Stig E. Bojesen, Sune F. Nielsen, Borge G. Nordestgaard, and the staff of the Copenhagen DNA laboratory, and Julie M. Cunningham, Sharon A. Windebank, Christopher A. Hilker, Jeffrey Meyer and the staff of Mayo Clinic Genotyping Core Facility

**Additional funding and acknowledgments from studies in PRACTICAL:**

Information of the PRACTICAL consortium can be found at <http://practical.icr.ac.uk/>

Aarhus

This study was supported by Innovation Fund Denmark, the Danish Cancer Society and The Velux Foundation (Veluxfonden). The Danish Cancer Biobank (DCB) is acknowledged for biological material.

AHS

This work was supported by the Intramural Research Program of the NIH, National Cancer Institute, Division of Cancer Epidemiology and Genetics (Z01CP010119).

ATBC

The ATBC Study is supported by the Intramural Research Program of the U.S. National Cancer Institute, National Institutes of Health, and by U.S. Public Health Service contract HHSN261201500005C from the National Cancer Institute, Department of Health and Human Services.

BioVU

The dataset(s) used for the analyses described were obtained from Vanderbilt University Medical Center’s BioVU which is supported by institutional funding and by the National Center for Research Resources, Grant UL1 RR024975-01 (which is now at the National Center for Advancing Translational Sciences, Grant 2 UL1 TR000445-06).

Canary PASS

PASS was supported by Canary Foundation and the National Cancer Institute's Early Detection Research Network (U01 CA086402)

CAPS & STHM1

The Department of Medical Epidemiology and Biostatistics, Karolinska Institute, Stockholm, Sweden was supported by the Cancer Risk Prediction Center (CRisP; www.crispcenter.org), a Linneus Centre (Contract ID 70867902) financed by the Swedish Research Council, Swedish Research Council (grant no K2010-70X-20430-04-3), the Swedish Cancer Foundation (grant no 09-0677), the Hedlund Foundation, the Soederberg Foundation, the Enqvist Foundation, ALF funds from the Stockholm County Council. Stiftelsen Johanna Hagstrand och Sigfrid Linner's Minne, Karlsson's Fund for urological and surgical research.	We thank and acknowledge all of the participants in the Stockholm-1 study. We thank Carin Cavalli-Bjoerkman and Ami Roennberg Karlsson for their dedicated work in the collection of data. Michael Broms is acknowledged for his skilful work with the databases. KI Biobank is acknowledged for handling the samples and for DNA extraction. Hans Wallinder at Aleris Medilab and Sven Gustafsson at Karolinska University Laboratory are thanked for their good cooperation in providing historical laboratory results.

CCI

This work was awarded by Prostate Cancer Canada and is proudly funded by the Movember Foundation - Grant # D2013-36.	The CCI group would like to thank David Murray, Razmik Mirzayans, and April Scott for their contribution to this work.

CHIPGECS

Orchid, Wuxi Second Hospital Research Funds, National Natural Science foundation of China for funding support to H Zhang (Grant No: 30671793 and 81072377), N Feng (Grant No: 81272831), SC Zhao (Grant No: 81072092 and 81328017), Liaoning Natural Science Foundation 2017, China, Item Number: 20170540536 to Zan Sun and Yong-Jie Lu.	This work was conducted on behalf of the CHIPGECS Consortia. We acknowledge the contribution of doctors, nurses and postgraduate research students at the CHIPGENCS sample collecting centers.

COH

SLN is partially supported by the Morris and Horowitz Families Endowed Professorship

CONOR

CONOR was supported by grants from the Nordic Cancer Union, the Swedish Cancer Society (2012/823) and the Swedish Research Council (2014/2269). The authors wish to acknowledge the services of CONOR, the contributing research centres delivering data to CONOR, and all the study participants.

COSM

COSM is funded by The Swedish Research Council (grant for the Swedish Infrastructure for Medical Population-based Life-course Environmental Research – SIMPLER), the Swedish Cancer Foundation.

CPCS1 / CPCS2

Department of Clinical Biochemistry, Herlev and Gentofte Hospital, Copenhagen University Hospital, Herlev Ringvej 75, DK-2730 Herlev, Denmark	We thank participants and staff of the Copenhagen General Population Study for their important contributions.

CPDR

Uniformed Services University for the Health Sciences HU0001-10-2-0002 (PI: David G. McLeod, MD)

EPIC

The coordination of EPIC was financially supported by the European Commission (DG-SANCO) and the International Agency for Research on Cancer. The national cohorts (that recruited male participants) are supported by Danish Cancer Society (Denmark); German Cancer Aid, German Cancer Research Center (DKFZ), Federal Ministry of Education and Research (BMBF), Deutsche Krebshilfe, Deutsches Krebsforschungszentrum and Federal Ministry of Education and Research (Germany); the Hellenic Health Foundation (Greece); Associazione Italiana per la Ricerca sul Cancro-AIRC-Italy and National Research Council (Italy); Dutch Ministry of Public Health, Welfare and Sports (VWS), Netherlands Cancer Registry (NKR), LK Research Funds, Dutch Prevention Funds, Dutch ZON (Zorg Onderzoek Nederland), World Cancer Research Fund (WCRF), Statistics Netherlands (The Netherlands); Health Research Fund (FIS), PI13/00061 to Granada; , PI13/01162 to EPIC-Murcia), Regional Governments of Andalucía, Asturias, Basque Country, Murcia and Navarra, ISCIII RETIC (RD06/0020) (Spain); Swedish Cancer Society, Swedish Research Council and County Councils of Skåne and Västerbotten (Sweden); Cancer Research UK (14136 to EPIC-Norfolk; C570/A16491 and C8221/A19170 to EPIC-Oxford), Medical Research Council (1000143 to EPIC-Norfolk, MR/M012190/1 to EPIC-Oxford) (United Kingdom). For information on how to submit an application for gaining access to EPIC data and/or biospecimens, please follow the instructions at http://epic.iarc.fr/access/index.php

EPICAP

The EPICAP study was supported by grants from Ligue Nationale Contre le Cancer; Institut National du Cancer (INCa); Fondation ARC; Fondation de France; Agence Nationale de sécurité sanitaire de l’alimentation, de l’environnement et du travail (ANSES); Ligue départementale du Val de Marne.	The EPICAP study group would like to thank all urologists, Antoinette Anger and Hasina Randrianasolo (study monitors), Anne-Laure Astolfi, Coline Bernard, Oriane Noyer, Marie-Hélène De Campo, Sandrine Margaroline, Louise N'Diaye, Sabine Perrier-Bonnet (Clinical Research nurses)

ERSPC

This study was supported by the DutchCancerSociety(KWF94-869,98-1657,2002-277,2006-3518, 2010-4800); The Netherlands Organisation for HealthResearch and Development (ZonMW-002822820,22000106,50-50110-98-311, 62300035), The Dutch Cancer Research Foundation(SWOP), and an unconditional grant from Beckman-Coulter-HybritechInc.

ESTHER

The ESTHER study was supported by a grant from the Baden Württemberg Ministry of Science, Research and Arts. The ESTHER group would like to thank Hartwig Ziegler, Sonja Wolf, Volker Hermann, Heiko Müller, Karina Dieffenbach, Katja Butterbach for valuable contributions to the study.

FHCRC

The FHCRC studies were supported by grants R01-CA056678, R01-CA082664, R01-CA092579, and K05-CA175147 from the US National Cancer Institute, National Institutes of Health, with additional support from the Fred Hutchinson Cancer Research Center (P30-CA015704). We thank all the men who participated in these studies.

Gene-PARE

The Gene-PARE study was supported by grants 1R01CA134444 from the U.S. National Institutes of Health, PC074201 and W81XWH-15-1-0680 from the Prostate Cancer Research Program of the Department of Defense and RSGT-05-200-01-CCE from the American Cancer Society. S.L.K. is supported by 1K07CA187546 from the U.S. National Cancer Institute.

HPFS

The Health Professionals Follow-up Study was supported by grants UM1CA167552, CA133891, CA141298, and P01CA055075. We are grateful to the participants and staff of the Physicians’ Health Study and Health Professionals Follow-Up Study for their valuable contributions, as well as the following state cancer registries for their help: AL, AZ, AR, CA, CO, CT, DE, FL, GA, ID, IL, IN, IA, KY, LA, ME, MD, MA, MI, NE, NH, NJ, NY, NC, ND, OH, OK, OR, PA, RI, SC, TN, TX, VA, WA, WY.

IMPACT

The IMPACT study was funded by The Ronald and Rita McAulay Foundation, CR-UK Project grant (C5047/A1232), Cancer Australia, AICR Netherlands A10-0227, Cancer Australia and Cancer Council Tasmania, NIHR, EU Framework 6, Cancer Councils of Victorial and South Australia, Philanthropic donation to Northshore University Health System. We acknowledge support from the National Institute for Health Research (NIHR) to the Biomedical Research Centre at The Institute of Cancer Research and Royal Marsden Foundation NHS Trust.	We acknowledge the IMPACT study steering committee, collaborating centres and participants.

IPO-Porto

The IPO-Porto study was funded by Fundação para a Ciência e a Tecnologia (FCT; UID/DTP/00776/2013 and PTDC/DTP-PIC/1308/2014) and FEDER (POCI-01-0145-FEDER-028245). MC (SFRH/BD/116557/2016) and MPS (SFRH/BD/132441/2017) are research fellows from FCT. We would like to express our gratitude to all patients and families who have participated in this study.

KARUPROSTATE

The Karuprostate study was supported by the the Frech National Health Directorate, the Association pour la Recherche sur le Cancer, la Ligue Nationale contre le Cancer, the French Agency for Environmental and Occupational Health Safety (ANSES) and by the Association pour la Recherche sur les Tumeurs de la Prostate. We would like to thank Séverine Ferdinand for valuable contributions to the study.

KULEUVEN. F.C. and S.J. are holders of grants from FWO Vlaanderen (G.0684.12N and G.0830.13N), the Belgian federal government (National Cancer Plan KPC_29_023), and a Concerted Research Action of the KU Leuven (GOA/15/017). TVDB is holder of a doctoral fellowship of the FWO.

LAAPC

This study was funded by grant R01CA84979 (to S.A. Ingles) from the National Cancer Institute, NIH.

Malaysia

The study was funded by the University Malaya High Impact Research Grant (HIR/MOHE/MED/35 to A.R). We thank all associates in the Urology Unit, University of Malaya, Cancer Research Malaysia (CRM) and the Malaysian Men's Health Initiative (MMHI).

MAYO

The Mayo group was supported by the US National Cancer Institute (R01CA72818)

MCC-Spain

The study was partially funded by the ""Accion Transversal del Cancer"", approved on the Spanish Ministry Council on the 11th October 2007, by the Instituto de Salud Carlos III-FEDER (PI08/1770, PI09/00773-Cantabria, PI11/01889-FEDER, PI12/00265, PI12/01270, PI12/00715, PI15/00069), by the Fundación Marqués de Valdecilla (API 10/09), by the Spanish Association Against Cancer (AECC) Scientific Foundation and by the Catalan Government DURSI grant 2009SGR1489. Samples: Biological samples were stored at the Parc de Salut MAR Biobank (MARBiobanc; Barcelona) which is supported by Instituto de Salud Carlos III FEDER (RD09/0076/00036). Also sample collection was supported by the Xarxa de Bancs de Tumors de Catalunya sponsored by Pla Director d'Oncologia de Catalunya (XBTC). ISGlobal is a member of the CERCA Programme, Generalitat de Catalunya. We acknowledge the contribution from Esther Gracia-Lavedan in preparing the data. We thank all the subjects who participated in the study and all MCC-Spain collaborators.

MCCS

Melbourne Collaborative Cohort Study (MCCS) cohort recruitment was funded by VicHealth and Cancer Council Victoria. The MCCS was further augmented by Australian National Health and Medical Research Council grants 209057, 396414 and 1074383 and by infrastructure provided by Cancer Council Victoria. Cases and their vital status were ascertained through the Victorian Cancer Registry and the Australian Institute of Health and Welfare, including the National Death Index and the Australian Cancer Database.

MD Anderson

Prostate Cancer Case-Control Studies at MD Anderson (MDA) supported by grants CA68578, ES007784, DAMD W81XWH-07-1-0645 and CA140388.

MEC

The MEC was supported by NIH grants CA063464, CA054281, CA098758, and CA164973.

MOFFITT

The Moffitt group was supported by the US National Cancer Institute (R01CA128813, PI: J.Y. Park).

NMHS

Funding for the Nashville Men's Health Study (NMHS) was provided by the National Institutes of Health Grant numbers: RO1CA121060

PCaP

The North Carolina - Louisiana Prostate Cancer Project (PCaP) and the Health Care Access and Prostate Cancer Treatment in North Carolina (HCaP-NC) study are carried out as collaborative studies supported by the Department of Defense contract DAMD 17-03-2-0052 and the American Cancer Society award RSGT-08-008-01-CPHPS, respectively. The authors thank the staff, advisory committees and research subjects participating in the PCaP study for their important contributions. We would like to acknowledge the UNC BioSpecimen Facility and the LSUHSC Pathology Lab for our DNA extractions, blood processing, storage, and sample disbursement (https://genome.unc.edu/bsp). The authors thank the staff, advisory committees and research subjects participating in the PCaP and HCaP-NC studies for their important contributions.

PCMUS

The PCMUS study was supported by the Bulgarian National Science Fund, Ministry of Education and Science (contract DOO-119/2009; DUNK01/2-2009; DFNI-B01/28/2012) with additional support from the Science Fund of Medical University - Sofia (contract 51/2009; 8I/2009; 28/2010; ).

PHS

The Physicians’ Health Study was supported by grants CA34944, CA40360, CA097193, HL26490 and HL34595. We are grateful to the participants and staff of the Physicians’ Health Study and Health Professionals Follow-Up Study for their valuable contributions, as well as the following state cancer registries for their help: AL, AZ, AR, CA, CO, CT, DE, FL, GA, ID, IL, IN, IA, KY, LA, ME, MD, MA, MI, NE, NH, NJ, NY, NC, ND, OH, OK, OR, PA, RI, SC, TN, TX, VA, WA, WY.

PLCO

This PLCO study was supported by the Intramural Research Program of the Division of Cancer Epidemiology and Genetics, National Cancer Institute, NIH. The authors thank Drs. Christine Berg and Philip Prorok, Division of Cancer Prevention at the National Cancer Institute, the screening center investigators and staff of the PLCO Cancer Screening Trial for their contributions to the PLCO Cancer Screening Trial. We thank Mr. Thomas Riley, Mr. Craig Williams, Mr. Matthew Moore, and Ms. Shannon Merkle at Information Management Services, Inc., for their management of the data and Ms. Barbara O’Brien and staff at Westat, Inc. for their contributions to the PLCO Cancer Screening Trial. We also thank the PLCO study participants for their contributions to making this study possible.

PRAGGA

PRAGGA was supported by Programa Grupos Emergentes, Cancer Genetics Unit, CHUVI Vigo Hospital, Instituto de Salud Carlos III, Spain. PRAGGA wishes to thank Victor Muñoz Garzón, Manuel Enguix Castelo, Sara Miranda Ponte, Carmen M Redondo, Manuel Calaza, Francisco Gude Sampedro, Joaquín González-Carreró and the staff of the Department of Pathology and Biobank of University Hospital Complex of Vigo, Instituto de Investigación Sanitaria Galicia Sur (IISGS), SERGAS, Vigo, Spain; Máximo Fraga, José Antúnez and the Biobank of University Hospital Complex of Santiago, Santiago de Compostela, Spain; and Maria Torres, Angel Carracedo and the Galician Foundation of Genomic Medicine.

PROCAP

PROCAP was supported by the Swedish Cancer Foundation (08-708, 09-0677).	We thank and acknowledge all of the participants in the PROCAP study. We thank Carin Cavalli-Björkman and Ami Rönnberg Karlsson for their dedicated work in the collection of data. Michael Broms is acknowledged for his skilful work with the databases. KI Biobank is acknowledged for handling the samples and for DNA extraction. We acknowledge The NPCR steering group: Pär Stattin (chair), Anders Widmark, Stefan Karlsson, Magnus Törnblom, Jan Adolfsson, Anna Bill-Axelson, Ove Andrén, David Robinson, Bill Pettersson, Jonas Hugosson, Jan-Erik Damber, Ola Bratt, Göran Ahlgren, Lars Egevad, and Roy Ehrnström.

PROFILE

We would like to acknowledge the support of the Ronald and Rita McAulay Foundation and Cancer Research UK. We also acknowledge support from the National Institute for Health Research (NIHR) to the Biomedical Research Centre at The Institute of Cancer Research and Royal Marsden Foundation NHS Trust	. We acknowledge the Profile study steering committee and participants.

PROGReSS

This research was supported by Spanish Instituto de Salud Carlos III (ISCIII) funding, an initiative of the Spanish Ministry of Economy and Innovation partially supported by European Regional Development FEDER Funds (INT15/00070, INT16/00154, INT17/00133; PI19/01424; PI16/00046; PI13/02030; PI10/00164), and through the Autonomous Government of Galicia (Consolidation and structuring program: IN607B) given to A.Vega. We would like to thank the patients for their contribution to the study

ProMPT & ProtecT

ProtecT funding: NIHR

ProMPT Funding*:* CRUK, NIHR, MRC, Cambridge Biomedical Research Centre

ProtecT would like to acknowledge the support of The University of Cambridge, Cancer Research UK. Cancer Research UK grants [C8197/A10123] and [C8197/A10865] supported the genotyping team. We would also like to acknowledge the support of the National Institute for Health Research which funds the Cambridge Bio-medical Research Centre, Cambridge, UK. We would also like to acknowledge the support of the National Cancer Research Prostate Cancer: Mechanisms of Progression and Treatment (PROMPT) collaborative (grant code G0500966/75466) which has funded tissue and urine collections in Cambridge. We are grateful to staff at the Welcome Trust Clinical Research Facility, Addenbrooke’s Clinical Research Centre, Cambridge, UK for their help in conducting the ProtecT study. We also acknowledge the support of the NIHR Cambridge Biomedical Research Centre, the NIR HTA (ProtecT grant) and the NCRI / MRC (ProMPT grant) for help with the bio-repository. The UK Department of Health funded the ProtecT study through the NIHR Health Technology Assessment Programme (projects 96/20/06, 96/20/99). The ProtecT trial and its linked ProMPT and CAP (The Cluster Randomized Trial of PSA Testing for Prostate Cancer) studies are supported by Department of Health, England; Cancer Research UK grant number C522/A8649, C11043/A4286, C18281/A8145, C18281/A11326, and C18281/A15064, Medical Research Council of England grant number G0500966, ID 75466 and The NCRI, UK. The epidemiological data for ProtecT were generated though funding from the Southwest National Health Service Research and Development. DNA extraction in ProtecT was supported by USA Dept of Defense award W81XWH-04-1-0280, Yorkshire Cancer Research and Cancer Research UK. The authors would like to acknowledge the contribution of all members of the ProtecT study research group. Richard Martin was supported by a Cancer Research UK Programme Grant (C18281/A19169) and the National Institute for Health Research Bristol Nutrition Biomedical Research Centre based at University Hospitals Bristol NHS Foundation Trust and the University of Bristol.

The views and opinions expressed therein are those of the authors and do not necessarily reflect those of the Department of Health of England. The bio-repository from ProtecT is supported by the NCRI (ProMPT) Prostate Cancer Collaborative and the Cambridge BMRC grant from NIHR.

We acknowledge support from the National Cancer Research Institute (National Institute of Health Research (NIHR) Collaborative Study: “Prostate Cancer: Mechanisms of Progression and Treatment (PROMPT)” (grant G0500966/75466).

We thank the National Institute for Health Research, Hutchison Whampoa Limited, the Human Research Tissue Bank (Addenbrooke’s Hospital), and Cancer Research UK. The authors would like to thank those men with prostate cancer and the subjects who have donated their time and their samples to the Cambridge Biorepository, which were used in this research. We also would like to acknowledge to support of the research staff in S4 who so carefully curated the samples and the follow-up data (Jo Burge, Marie Corcoran, Anne George, and Sara Stearn).

PROtEuS

PROtEuS was supported financially through grants from the Canadian Cancer Society [13149, 19500, 19864, 19865] and the Cancer Research Society, in partnership with the Ministère de l'enseignement supérieur, de la recherche, de la science et de la technologie du Québec, and the Fonds de la recherche du Québec - Santé. PROtEuS would like to thank its collaborators and research personnel, and the urologists involved in subjects recruitment. We also wish to acknowledge the special contribution made by Ann Hsing and Anand Chokkalingam to the conception of the genetic component of PROtEuS.

QLD

The QLD research is supported by The National Health and Medical Research Council (NHMRC) Australia Project Grants [390130, 1009458] and NHMRC Career Development Fellowship, Cancer Australia PdCCRS and Cancer Council Queensland funding to J Batra. The QLD team would like to acknowledge and sincerely thank the urologists, pathologists, data managers and patient participants who have generously and altruistically supported the QLD cohort.

RAPPER

RAPPER is supported by Cancer Research UK [C1094/A11728; C1094/A18504] and Experimental Cancer Medicine Centre funding [C1467/A7286], and the NIHR Manchester Biomedical Research Centre. The RAPPER group thank Dr. Holly Summersgill for project management.

SABOR

The SABOR research is supported by NIH/NCI Early Detection Research Network, grant U01 CA0866402-18. Also supported by the Cancer Center Support Grant to the Mays Cancer Center from the National Cancer Institute (US) P30 CA054174

SCCS

SCCS is funded by NIH grant R01 CA092447, and SCCS sample preparation was conducted at the Epidemiology Biospecimen Core Lab that is supported in part by the Vanderbilt-Ingram Cancer Center (P30 CA68485). Data on SCCS cancer cases used in this publication were provided by the Alabama Statewide Cancer Registry; Kentucky Cancer Registry, Lexington, KY; Tennessee Department of Health, Office of Cancer Surveillance; Florida Cancer Data System; North Carolina Central Cancer Registry, North Carolina Division of Public Health; Georgia Comprehensive Cancer Registry; Louisiana Tumor Registry; Mississippi Cancer Registry; South Carolina Central Cancer Registry; Virginia Department of Health, Virginia Cancer Registry; Arkansas Department of Health, Cancer Registry, 4815 W. Markham, Little Rock, AR 72205. The Arkansas Central Cancer Registry is fully funded by a grant from National Program of Cancer Registries, Centers for Disease Control and Prevention (CDC). Data on SCCS cancer cases from

**PRACTICAL Consortium Funding & Acknowledgements**

Genotyping of the OncoArray was funded by the US National Institutes of Health (NIH) [U19 CA 148537 for ELucidating Loci Involved in Prostate cancer SuscEptibility (ELLIPSE) project and X01HG007492 to the Center for Inherited Disease Research (CIDR) under contract number HHSN268201200008I]. Additional analytic support was provided by NIH NCI U01 CA188392 (PI: Schumacher).

The PRACTICAL consortium was supported by Cancer Research UK Grants C5047/A7357, C1287/A10118, C1287/A16563, C5047/A3354, C5047/A10692, C16913/A6135, European Commission's Seventh Framework Programme grant agreement n° 223175 (HEALTH-F2-2009-223175), and The National Institute of Health (NIH) Cancer Post-Cancer GWAS initiative grant: No. 1 U19 CA 148537-01 (the GAME-ON initiative).

We would also like to thank the following for funding support: The Institute of Cancer Research and The Everyman Campaign, The Prostate Cancer Research Foundation, Prostate Research Campaign UK (now PCUK), The Orchid Cancer Appeal, Rosetrees Trust, The National Cancer Research Network UK, The National Cancer Research Institute (NCRI) UK. We are grateful for support of NIHR funding to the NIHR Biomedical Research Centre at The Institute of Cancer Research and The Royal Marsden NHS Foundation Trust.

This study would not have been possible without the contributions of the following:

Coordination team, bioinformatician and genotyping centres:

Genotyping at CCGE, Cambridge: Caroline Baines and Don Conroy

Additional funding and acknowledgments from studies in PRACTICAL:

Information of the PRACTICAL consortium can be found at

<http://practical.icr.ac.uk/>

Aarhus

This study was supported by Innovation Fund Denmark, the Danish Cancer Society and The Velux Foundation (Veluxfonden).

The Danish Cancer Biobank (DCB) is acknowledged for biological material.

AHS

This work was supported by the Intramural Research Program of the NIH, National Cancer Institute, Division of Cancer Epidemiology and Genetics (Z01CP010119).

ATBC

The ATBC Study is supported by the Intramural Research Program of the U.S. National Cancer Institute, National Institutes of Health, Department of Health and Human Services.

BioVU

The dataset(s) used for the analyses described were obtained from Vanderbilt University Medical Center’s BioVU which is supported by institutional funding and by the National Center for Research Resources, Grant UL1 RR024975-01 (which is now at the National Center for Advancing Translational Sciences, Grant 2 UL1 TR000445-06).

Canary PASS

PASS was supported by Canary Foundation and the National Cancer Institute's Early Detection Research Network )U01 CA086402)

CCI

This work was awarded by Prostate Cancer Canada and is proudly funded by the Movember Foundation - Grant # D2013-36.

The CCI group would like to thank David Murray, Razmik Mirzayans, and April Scott for their contribution to this work.

CHIPGECS

Orchid, Wuxi Second Hospital Research Funds, National Natural Science foundation of China for funding support to H Zhang (Grant No: 30671793 and 81072377), N Feng (Grant No: 81272831), SC Zhao (Grant No: 81072092 and 81328017), Liaoning Natural Science Foundation 2017, China, Item Number: 20170540536 to Zan Sun Sun and Yong-Jie Lu.

This work was conducted on behalf of the CHIPGECS Consortia. We acknowledge the contribution of doctors, nurses and postgraduate research students at the CHIPGENCS sample collecting centers.

COH

SLN is partially supported by the Morris and Horowitz Families Endowed Professorship

COSM

COSM is funded by The Swedish Research Council (grant for the Swedish Infrastructure for Medical Population-based Life-course Environmental Research – SIMPLER), the Swedish Cancer Foundation.

CPCS1 / CPCS2

Department of Clinical Biochemistry, Herlev and Gentofte Hospital, Copenhagen University Hospital, Herlev Ringvej 75, DK-2730 Herlev, Denmark

We thank participants and staff of the Copenhagen General Population Study for their important contributions.

CPDR

Uniformed Services University for the Health Sciences HU0001-10-2-0002 (PI: David G. McLeod, MD)

EPIC

The coordination of EPIC was financially supported by the European Commission (DG-SANCO) and the International Agency for Research on Cancer. The national cohorts (that recruited male participants) are supported by Danish Cancer Society (Denmark); German Cancer Aid, German Cancer Research Center (DKFZ), Federal Ministry of Education and Research (BMBF), Deutsche Krebshilfe, Deutsches Krebsforschungszentrum and Federal Ministry of Education and Research (Germany); the Hellenic Health Foundation (Greece); Associazione Italiana per la Ricerca sul Cancro-AIRC-Italy and National Research Council (Italy); Dutch Ministry of Public Health, Welfare and Sports (VWS), Netherlands Cancer Registry (NKR), LK Research Funds, Dutch Prevention Funds, Dutch ZON (Zorg Onderzoek Nederland), World Cancer Research Fund (WCRF), Statistics Netherlands (The Netherlands); Health Research Fund (FIS), PI13/00061 to Granada; , PI13/01162 to EPIC-Murcia), Regional Governments of Andalucía, Asturias, Basque Country, Murcia and Navarra, ISCIII RETIC (RD06/0020) (Spain); Swedish Cancer Society, Swedish Research Council and County Councils of Skåne and Västerbotten (Sweden); Cancer Research UK (14136 to EPIC-Norfolk; C8221/A29017 to EPIC-Oxford), Medical Research Council

(1000143 to EPIC-Norfolk, MR/M012190/1 to EPIC-Oxford) (United Kingdom). For information on how to submit an application for gaining access to EPIC data and/or biospecimens, please follow the instructions at http://epic.iarc.fr/access/index.php

EPICAP

The EPICAP study was supported by grants from Ligue Nationale Contre le Cancer; Institut National du Cancer (INCa); Fondation ARC; Fondation de France; Agence Nationale de sécurité sanitaire de l’alimentation, de l’environnement et du travail (ANSES); Ligue départementale du Val de Marne.

The EPICAP study group would like to thank all urologists, Antoinette Anger and Hasina Randrianasolo (study monitors), Anne-Laure Astolfi, Coline Bernard, Oriane Noyer, Marie-Hélène De Campo, Sandrine Margaroline, Louise N'Diaye, Sabine Perrier-Bonnet (Clinical Research nurses)

ERSPC

This study was supported by the DutchCancerSociety(KWF94-869,98-1657,2002-277,2006-3518, 2010-4800); The Netherlands Organisation for HealthResearch and Development (ZonMW-002822820,22000106,50-50110-98-311, 62300035), The Dutch Cancer Research Foundation(SWOP), and an unconditional grant from Beckman-Coulter-HybritechInc.

ESTHER

The ESTHER study was supported by a grant from the Baden Württemberg Ministry of Science, Research and Arts.

The ESTHER group would like to thank Hartwig Ziegler, Sonja Wolf, Volker Hermann, Heiko Müller, Karina Dieffenbach, Katja Butterbach for valuable contributions to the study.

FHCRC

The FHCRC studies were supported by grants R01-CA080122, R01-CA056678, R01-CA082664, and R01-CA092579, and K05-CA175147 from the US National Cancer Institute, National Institutes of Health, with additional support from the Fred Hutchinson Cancer Research Center (P30-CA015704).

We thank all the individuals who participated in these studies.

Gene-PARE

The Gene-PARE study was supported by grants 1R01CA134444 from the U.S. National Institutes of Health , PC074201 and W81XWH-15-1-0680 from the Prostate Cancer Research Program of the Department of Defense and RSGT-05-200-01-CCE from the American Cancer Society. S.L.K. is supported by 1K07CA187546 from the U.S. National Cancer Institute.

HPFS

The Health Professionals Follow-up Study was supported by grants UM1CA167552, CA133891, CA141298, and P01CA055075.

We are grateful to the participants and staff of the Physicians’ Health Study and Health Professionals Follow-Up Study for their valuable contributions, as well as the following state cancer registries for their help: AL, AZ, AR, CA, CO, CT, DE, FL, GA, ID, IL, IN, IA, KY, LA, ME, MD, MA, MI, NE, NH, NJ, NY, NC, ND, OH, OK, OR, PA, RI, SC, TN, TX, VA, WA, WY.

IMPACT

The IMPACT study was funded by The Ronald and Rita McAulay Foundation, CR-UK Project grant (C5047/A1232), Cancer Australia, AICR Netherlands A10-0227, Cancer Australia and Cancer Council Tasmania, NIHR, EU Framework 6, Cancer Councils of Victorial and South Australia, Philanthropic donation to Northshore University Health System. We acknowledge support from the National Institute for Health Research (NIHR) to the Biomedical Research Centre at The Institute of Cancer Research and Royal Marsden Foundation NHS Trust.

We acknowledge the IMPACT study steering committee, collaborating centres and participants.

IPO-Porto

The IPO-Porto study was funded by Fundação para a Ciência e a Tecnologia (FCT; UID/DTP/00776/2013 and PTDC/DTP-PIC/1308/2014) and FEDER (POCI-01-0145-FEDER-028245). MC (SFRH/BD/116557/2016) and MPS (SFRH/BD/132441/2017) are research fellows from FCT.

We would like to express our gratitude to all patients and families who have participated in this study.

KARUPROSTATE

The Karuprostate study was supported by the French National Health Directorate, the Association pour la Recherche sur le Cancer, la Ligue Nationale contre le Cancer, the French Agency for Environmental and Occupational Health Safety (ANSES) and by the Association pour la Recherche sur les Tumeurs de la Prostate. We would like to thank Séverine Ferdinand for valuable contributions to the study.

KULEUVEN

F.C. and S.J. are holders of grants from FWO Vlaanderen (G.0684.12N and G.0830.13N), the Belgian federal government (National Cancer Plan KPC_29_023), and a Concerted Research Action of the KU Leuven (GOA/15/017). TVDB is holder of a doctoral fellowship of the FWO.

LAAPC

This study was funded by grant R01CA84979 (to S.A. Ingles) from the National Cancer Institute, NIH.

Malaysia

The study was funded by the University Malaya High Impact Research Grant (HIR/MOHE/MED/35 to A.R).

We thank all associates in the Urology Unit, University of Malaya, Cancer Research Malaysia (CRM) and the Malaysian Men's Health Initiative (MMHI).

MCC-Spain

The study was partially funded by the ""Accion Transversal del Cancer"", approved on the Spanish Ministry Council on the 11th October 2007, by the Instituto de Salud Carlos III-FEDER (PI08/1770, PI09/00773-Cantabria, PI11/01889-FEDER, PI12/00265, PI12/01270, PI12/00715, PI15/00069), by the Fundación Marqués de Valdecilla (API 10/09), by the Spanish Association Against Cancer (AECC) Scientific Foundation and by the Catalan Government DURSI grant 2009SGR1489. Samples: Biological samples were stored at the Parc de Salut MAR Biobank (MARBiobanc; Barcelona) which is supported by Instituto de Salud Carlos III FEDER (RD09/0076/00036). Also sample collection was supported by the Xarxa de Bancs de Tumors de Catalunya sponsored by Pla Director d'Oncologia de Catalunya (XBTC). ISGlobal acknowledges support from the Spanish Ministry of Science and Innovation through the “Centro de Excelencia Severo Ochoa 2019-2023” Program (CEX2018-000806-S), and support from the Generalitat de Catalunya through the CERCA Program.

We acknowledge the contribution from Esther Gracia-Lavedan in preparing the data. We thank all the subjects who participated in the study and all MCC-Spain collaborators.

MCCS

Melbourne Collaborative Cohort Study (MCCS) cohort recruitment was funded by VicHealth and Cancer Council Victoria. The MCCS was further augmented by Australian National Health and Medical Research Council grants 209057, 396414 and 1074383 and by infrastructure provided by Cancer Council Victoria. Cases and their vital status were ascertained through the Victorian Cancer Registry and the Australian Institute of Health and Welfare, including the National Death Index and the Australian Cancer Database.

MD Anderson

Prostate Cancer Case-Control Studies at MD Anderson (MDA) supported by grants CA68578, ES007784, DAMD W81XWH-07-1-0645 and CA140388.

MEC

The MEC was supported by NIH grants CA063464, CA054281, CA098758, and CA164973.

MOFFITT

The Moffitt group was supported by the US National Cancer Institute (R01CA128813, PI: J.Y. Park).

NMHS

Funding for the Nashville Men's Health Study (NMHS) was provided by the National Institutes of Health Grant numbers: RO1CA121060

Oslo

CONOR was supported by grants from the Nordic Cancer Union, the Swedish Cancer Society (2012/823) and the Swedish Research Council (2014/2269).

The authors wish to acknowledge the services of CONOR, the contributing research centres delivering data to CONOR, and all the study participants.

PCaP

The North Carolina - Louisiana Prostate Cancer Project (PCaP) and the Health Care Access and Prostate Cancer Treatment in North Carolina (HCaP-NC) study are carried out as collaborative studies supported by the Department of Defense contract DAMD 17-03-2-0052 and the American Cancer Society award RSGT-08-008-01-CPHPS, respectively.

We would like to acknowledge the UNC BioSpecimen Facility and the LSUHSC Pathology Lab for our DNA extractions, blood processing, storage and sample disbursement (<https://genome.unc.edu/bsp>).

The Health Care Access and Prostate Cancer Treatment in North Carolina (HCaP-NC) study is carried out as a collaborative study supported by the American Cancer Society award RSGT-08-008-01-CPHPS. The authors thank the staff, advisory committees and research subjects participating in the PCaP and HCaP-NC studies for their important contributions.

PCMUS

The PCMUS study was supported by the Bulgarian National Science Fund, Ministry of Education and Science (contract DOO-119/2009; DUNK01/2-2009; DFNI-B01/28/2012) with additional support from the Science Fund of Medical University - Sofia (contract 51/2009; 8I/2009; 28/2010; ).

PHS

The Physicians’ Health Study was supported by grants CA34944, CA40360, CA097193, HL26490 and HL34595.

We are grateful to the participants and staff of the Physicians’ Health Study and Health Professionals Follow-Up Study for their valuable contributions, as well as the following state cancer registries for their help: AL, AZ, AR, CA, CO, CT, DE, FL, GA, ID, IL, IN, IA, KY, LA, ME, MD, MA, MI, NE, NH, NJ, NY, NC, ND, OH, OK, OR, PA, RI, SC, TN, TX, VA, WA, WY.

PLCO

This PLCO study was supported by the Intramural Research Program of the Division of Cancer Epidemiology and Genetics, National Cancer Institute, NIH

The authors thank Drs. Christine Berg and Philip Prorok, Division of Cancer Prevention at the National Cancer Institute, the screening center investigators and staff of the PLCO Cancer Screening Trial for their contributions to the PLCO Cancer Screening Trial. We thank Mr. Thomas Riley, Mr. Craig Williams, Mr. Matthew Moore, and Ms. Shannon Merkle at Information Management Services, Inc., for their management of the data and Ms. Barbara O’Brien and staff at Westat, Inc. for their contributions to the PLCO Cancer Screening Trial. We also thank the PLCO study participants for their contributions to making this study possible.

PRAGGA

PRAGGA was supported by Programa Grupos Emergentes, Cancer Genetics Unit, CHUVI Vigo Hospital, Instituto de Salud Carlos III, Spain.

PRAGGA wishes to thank Victor Muñoz Garzón, Manuel Enguix Castelo, Sara Miranda Ponte, Carmen M Redondo, Manuel Calaza, Francisco Gude Sampedro, Joaquín González-Carreró and the staff of the Department of Pathology and Biobank of University Hospital Complex of Vigo, Instituto de Investigación Sanitaria Galicia Sur (IISGS), SERGAS, Vigo, Spain; Máximo Fraga, José Antúnez and the Biobank of University Hospital Complex of Santiago, Santiago de Compostela, Spain; and Maria Torres, Angel Carracedo and the Galician Foundation of Genomic Medicine.

PROCAP

PROCAP was supported by the Swedish Cancer Foundation (08-708, 09-0677).

We thank and acknowledge all of the participants in the PROCAP study. We thank Carin Cavalli-Björkman and Ami Rönnberg Karlsson for their dedicated work in the collection of data. Michael Broms is acknowledged for his skilful work with the databases. KI Biobank is acknowledged for handling the samples and for DNA extraction. We acknowledge The NPCR steering group: Pär Stattin (chair), Anders Widmark, Stefan Karlsson, Magnus Törnblom, Jan Adolfsson, Anna Bill-Axelson, Ove Andrén, David Robinson, Bill Pettersson, Jonas Hugosson, Jan-Erik Damber, Ola Bratt, Göran Ahlgren, Lars Egevad, and Roy Ehrnström.

PROFILE

We would like to acknowledge the support of the Ronald and Rita McAulay Foundation and Cancer Research UK. We also acknowledge support from the National Institute for Health Research (NIHR) to the Biomedical Research Centre at The Institute of Cancer Research and Royal Marsden Foundation NHS Trust

We acknowledge the Profile study steering committee and participants.

PROGReSS

This research was supported by Spanish Instituto de Salud Carlos III (ISCIII) funding, an initiative of the Spanish Ministry of Economy and Innovation partially supported by European Regional Development FEDER Funds (INT15/00070, INT16/00154, INT17/00133; PI19/01424; PI16/00046; PI13/02030; PI10/00164), and through the Autonomous Government of Galicia (Consolidation and structuring program: IN607B) given to A.Vega.

We would like to thank the patients for their contribution to the study

ProMPT / ProtecT

CRUK, NIHR, MRC, Cambridge Biomedical Research Centre

"ProtecT would like to acknowledge the support of The University of Cambridge, Cancer Research UK. Cancer Research UK grants [C8197/A10123] and [C8197/A10865] supported the genotyping team. We would also like to acknowledge the support of the National Institute for Health Research which funds the Cambridge Bio-medical Research Centre, Cambridge, UK. We would also like to acknowledge the support of the National Cancer Research Prostate Cancer: Mechanisms of Progression and Treatment (PROMPT) collaborative (grant code G0500966/75466) which has funded tissue and urine collections in Cambridge. We are grateful to staff at the Welcome Trust Clinical Research Facility, Addenbrooke’s Clinical Research Centre, Cambridge, UK for their help in conducting the ProtecT study. We also acknowledge the support of the NIHR Cambridge Biomedical Research Centre, the NIR HTA (ProtecT grant) and the NCRI / MRC (ProMPT grant) for help with the bio-repository. The UK Department of Health funded the ProtecT study through the NIHR Health Technology Assessment Programme (projects 96/20/06, 96/20/99). The ProtecT trial and its linked ProMPT and CAP (The Cluster Randomized Trial of PSA Testing for Prostate Cancer) studies are supported by Department of Health, England; Cancer Research UK grant number C522/A8649, C11043/A4286, C18281/A8145, C18281/A11326, and C18281/A15064, Medical Research Council of England grant number G0500966, ID 75466 and The NCRI, UK. The epidemiological data for ProtecT were generated though funding from the Southwest National Health Service Research and Development. DNA extraction in ProtecT was supported by USA Dept of Defense award W81XWH-04-1-0280, Yorkshire Cancer Research and Cancer Research UK. The authors would like to acknowledge the contribution of all members of the ProtecT study research group.

Richard Martin was supported by a Cancer Research UK Programme Grant (C18281/A19169) and the National Institute for Health Research Bristol Nutrition Biomedical Research Centre based at University Hospitals Bristol NHS Foundation Trust and the University of Bristol.

The views and opinions expressed therein are those of the authors and do not necessarily reflect those of the Department of Health of England. The bio-repository from ProtecT is supported by the NCRI (ProMPT) Prostate Cancer Collaborative and the Cambridge BMRC grant from NIHR.

We acknowledge support from the National Cancer Research Institute (National Institute of Health Research (NIHR) Collaborative Study: “Prostate Cancer: Mechanisms of Progression and Treatment (PROMPT)” (grant G0500966/75466).

We thank the National Institute for Health Research, Hutchison Whampoa Limited, the Human Research Tissue Bank (Addenbrooke’s Hospital), and Cancer Research UK. The authors would like to thank those men with prostate cancer and the subjects who have donated their time and their samples to the Cambridge Biorepository, which were used in this research. We also would like to acknowledge to support of the research staff in S4 who so carefully curated the samples and the follow-up data (Jo Burge, Marie Corcoran, Anne George, and Sara Stearn)."

PROtEuS

PROtEuS was supported financially through grants from the Canadian Cancer Society [13149, 19500, 19864, 19865] and the Cancer Research Society, in partnership with the Ministère de l'enseignement supérieur, de la recherche, de la science et de la technologie du Québec, and the Fonds de la recherche du Québec - Santé.

PROtEuS would like to thank its collaborators and research personnel, and the urologists involved in subjects recruitment. We also wish to acknowledge the special contribution made by Ann Hsing and Anand Chokkalingam to the conception of the genetic component of PROtEuS.

QLD

The QLD research is supported by The National Health and Medical Research Council (NHMRC) Australia Project Grants [390130, 1009458] and NHMRC Career Development Fellowship, Cancer Australia PdCCRS and Cancer Council Queensland funding to J Batra

The QLD team would like to acknowledge and sincerely thank the urologists, pathologists, data managers and patient participants who have generously and altruistically supported the QLD cohort.

RAPPER

RAPPER is supported by Cancer Research UK [C1094/A11728; C1094/A18504] and Experimental Cancer Medicine Centre funding [C1467/A7286], and the NIHR Manchester Biomedical Research Centre.

The RAPPER group thank Dr. Holly Summersgill for project management.

SABOR

The SABOR research is supported by NIH/NCI Early Detection Research Network, grant U01 CA0866402-18. Also supported by the Cancer Center Support Grant to the Mays Cancer Center from the National Cancer Institute (US) P30 CA054174

SCCS

SCCS is funded by NIH grant R01 CA092447, and SCCS sample preparation was conducted at the Epidemiology Biospecimen Core Lab that is supported in part by the Vanderbilt-Ingram Cancer Center (P30 CA68485). Data on SCCS cancer cases used in this publication were provided by the Alabama Statewide Cancer Registry; Kentucky Cancer Registry, Lexington, KY; Tennessee Department of Health, Office of Cancer Surveillance; Florida Cancer Data System; North Carolina Central Cancer Registry, North Carolina Division of Public Health; Georgia Comprehensive Cancer Registry; Louisiana Tumor Registry; Mississippi Cancer Registry; South Carolina Central Cancer Registry; Virginia Department of Health, Virginia Cancer Registry; Arkansas Department of Health, Cancer Registry, 4815 W. Markham, Little Rock, AR 72205. The Arkansas Central Cancer Registry is fully funded by a grant from National Program of Cancer Registries, Centers for Disease Control and Prevention (CDC). Data on SCCS cancer cases from Mississippi were collected by the Mississippi Cancer Registry which participates in the National Program of Cancer Registries (NPCR) of the Centers for Disease Control and Prevention (CDC). The contents of this publication are solely the responsibility of the authors and do not necessarily represent the official views of the CDC or the Mississippi Cancer Registry.

SCPCS

SCPCS is funded by CDC grant S1135-19/19, and SCPCS sample preparation was conducted at the Epidemiology Biospecimen Core Lab that is supported in part by the Vanderbilt-Ingram Cancer Center (P30 CA68485).

SEARCH

SEARCH is funded by a programme grant from Cancer Research UK [C490/A10124] and supported by the UK National Institute for Health Research Biomedical Research Centre at the University of Cambridge. The University of Cambridge has received salary support in respect of PP from the NHS in the East of England through the Clinical Academic Reserve.

SFPCS

SFPCS was funded by California Cancer Research Fund grant 99-00527V-10182

SNP_Prostate_Ghent

The study was supported by the National Cancer Plan, financed by the Federal Office of Health and Social Affairs, Belgium.

SPAG

Manchester Cancer Research Centre and MRC – Medical Research Council for PhD studentship work, MUG - Male Uprising in Guernsey, Hope for Guernsey and Wessex Medical Research for research support

STHM2

STHM2 was supported by grants from The Strategic Research Programme on Cancer (StratCan), Karolinska Institutet; the Linné Centre for Breast and Prostate Cancer (CRISP, number 70867901), Karolinska Institutet; The Swedish Research Council (number K2010-70X-20430-04-3) and The Swedish Cancer Society (numbers 11-0287 and 11-0624); Stiftelsen Johanna Hagstrand och Sigfrid Linnérs minne; Swedish Council for Working Life and Social Research (FAS), number 2012-0073

The authors acknowledge the Karolinska University Laboratory, Aleris Medilab, Unilabs and the Regional Prostate Cancer Registry for performing analyses and help to retrieve data. Carin Cavalli–Björkman and Britt-Marie Hune for their enthusiastic work as research nurses. Astrid Björklund for skilful data management. We wish to thank the BBMRI.se biobank facility at Karolinska Institutet for biobank services.

SWOG-PCPT / SWOG-SELECT

PCPT / SELECT is funded by Public Health Service grants U10CA37429 and 5UM1CA182883 from the National Cancer Institute.

The authors thank the site investigators and staff and, most importantly, the participants from PCPT who donated their time to this trial.

TAMPERE

The Tampere (Finland) study was supported by the Academy of Finland (251074), The Finnish Cancer Organisations, Sigrid Juselius Foundation, and the Competitive Research Funding of the Tampere University Hospital (X51003). The PSA screening samples were collected by the Finnish part of ERSPC (European Study of Screening for Prostate Cancer).

TAMPERE would like to thank Riina Liikanen, Liisa Maeaettaenen and Kirsi Talala for their work on samples and databases.

Toronto

Prostate Cancer Canada Movember Discovery Grant (D2013-17) to RJH; Canadian Cancer Society Research Institute Career Development Award in Cancer Prevention (2013-702108) to RJH

UKGPCS

UKGPCS would also like to thank the following for funding support: The Institute of Cancer Research and The Everyman Campaign, The Prostate Cancer Research Foundation, Prostate Research Campaign UK (now Prostate Action), The Orchid Cancer Appeal, The National Cancer Research Network UK, The National Cancer Research Institute (NCRI) UK. We are grateful for support of NIHR funding to the NIHR Biomedical Research Centre at The Institute of Cancer Research and The Royal Marsden NHS Foundation Trust. UKGPCS should also like to acknowledge the NCRN nurses, data managers and Consultants for their work in the UKGPCS.

UKGPCS would like to thank all urologists and other persons involved in the planning, coordination, and data collection of the study. KM and AL were in part supported from the NIHR Manchester Biomedical Research Centre

ULM

The Ulm group received funds from the German Cancer Aid (Deutsche Krebshilfe).

WUGS

WUGS would like to thank the following for funding support: The Anthony DeNovi Fund, the Donald C. McGraw Foundation, and the St. Louis Men’s Group Against Cancer.
